# A multi-level investigation of the genetic relationship between endometriosis and ovarian cancer histotypes

**DOI:** 10.1101/2021.06.28.21259290

**Authors:** Sally Mortlock, Rosario I. Corona, Pik Fang Kho, Paul Pharoah, Ji-Heui Seo, Matthew L. Freedman, Simon A. Gayther, Matthew T. Siedhoff, Peter A.W. Rogers, Ronald Leuchter, Christine S. Walsh, Ilana Cass, Beth Y. Karlan, B.J. Rimel, Ovarian Cancer Association Consortium, International Endometriosis Genetics Consortium, Grant W. Montgomery, Kate Lawrenson, Siddhartha P. Kar

**Author notes:** These authors jointly supervised this study and are co-last authors (Kate Lawrenson, Siddhartha P. Kar).

## Abstract

Endometriosis is associated with increased risk of epithelial ovarian cancers (EOCs). Data from large endometriosis and EOC genome-wide association meta-analyses were used to estimate the genetic correlation and evaluate the causal relationship between genetic liability to endometriosis and major EOC histotypes, and to identify shared susceptibility loci. We estimated a significant genetic correlation (r_g_) between endometriosis and clear cell (r_g_=0.71), endometrioid (r_g_=0.48) and high-grade serous (r_g_=0.19) ovarian cancer, all supported by Mendelian randomization analyses. A bivariate meta-analysis identified 28 loci associated with endometriosis and EOC, including two novel risk loci, and 19 with evidence for a single underlying causal variant. Differences in the shared risk suggest different underlying pathways may contribute to the relationship between endometriosis and the different histotypes. Functional annotation using transcriptomic and epigenomic profiles of relevant tissues/cells highlighted several target genes. This comprehensive analysis reveals profound genetic overlap between endometriosis and EOC histotypes with valuable genomic targets for understanding the biological mechanisms linking the diseases.

## Introduction

Endometriosis is a chronic gynaecological disease affecting up to 12% of reproductive-age women (Giudice, 2010; Rowlands *et al*, 2021; Treloar *et al*, 2002). The disease is characterised by the presence of endometriotic lesions outside the uterus and is associated with pelvic pain and subfertility (Giudice, 2010). Lesions are frequently categorized according to lesion location and depth of infiltration into the surrounding tissue and include superficial peritoneal lesions, deep infiltrating disease and cysts (endometriomas), most commonly found on the ovary (American Society for Reproductive, 1997). While endometriosis is a benign condition, it shares features with cancer including metastatic-like behaviour, tissue invasion, proliferation, angiogenesis and decreased apoptosis. Large epidemiologic studies have reproducibly shown that women with endometriosis have increased risk of epithelial ovarian cancer (standardized incidence ratio=1.8-8.95), although the absolute risks are quite small, and there is currently no way to predict which endometriosis patients are most likely to develop ovarian cancer later in life (Brilhante *et al*, 2017; Kok *et al*, 2015; Zafrakas *et al*, 2014). Somatic mutations shared between benign endometriotic lesions and adjacent tumours suggest these lesions are cellular precursors to endometriosis-associated ovarian cancers. Examples include loss of function mutations in *ARID1A* and gain of function mutations in *PIK3CA* (Anglesio *et al*, 2015; Jones *et al*, 2010; Wiegand *et al*, 2010b, a).

Ovarian cancer is the deadliest gynaecologic cancer. Fewer than 50% of women survive beyond 5 years after diagnosis due to the rapid development of chemoresistance and the absence of effective early detection strategies. Research is needed to advance understanding of disease aetiology, identify risk factors and develop early detection methods and effective targeted therapies. The major histological subtypes of EOC include high-grade serous (HGSOC), low-grade serous (LGSOC), mucinous (MOC), endometrioid (ENOC) and clear cell (CCOC) EOC. Borderline tumors of low malignant potential also exist, most typically with serous (LMPSOC) or mucinous differentiation. CCOC and ENOC are the two histotypes most strongly and reproducibly associated with endometriosis (Brinton *et al*, 2005; Pearce *et al*, 2012; Rossing *et al*, 2008). Concurrent endometriosis is observed in 21-51% patients with CCOC and 23-43% women with ENOC (Jimbo *et al*, 1997; Stamp *et al*, 2016; Vercellini *et al*, 1993). These histotype-associations are supported by observational study data from 7,911 women with invasive EOC in the Ovarian Cancer Association Consortium that showed a significant association between history of endometriosis and specific histological subtypes of ovarian cancer including CCOC (odds ratio (OR)=3.05), ENOC (OR=2.04) and, to a lesser extent, LGSOC (OR=2.11)(Pearce *et al*., 2012).

Recent genome-wide association studies (GWASs) have provided strong evidence for a genetic contribution to risk of both endometriosis and ovarian cancer. A total of 19 independent genomic regions have been associated with endometriosis risk and 34 have been associated with different histotypes of EOC (Phelan *et al*, 2017; Sapkota *et al*, 2017). Germline variants may also contribute to increased risk of developing both diseases. Lu *et al* (2015) used genetic data from 84,000 SNPs genotyped in EOC (10,065 cases) and endometriosis (3,194 cases) cohorts to estimate the genetic correlation between the diseases and found a strong significant genetic correlation between endometriosis and CCOC (0.51), ENOC (0.48), and LGSOC (0.40) and smaller correlation with HGSOC (0.25). This study, however, was limited by the relatively small number of SNPs and sample numbers and did not provide evidence for a causal relationship between genetic liability to endometriosis and EOC risk or shared risk loci. The aim of this present study was to use data from newer and larger endometriosis (14,949 cases/190,715 controls) and ovarian cancer (25,509 cases/40,941 controls) GWAS meta-analyses to estimate the genetic correlation and causal relationship between endometriosis and ovarian cancer histotypes and to identify shared genetic susceptibility loci, candidate target genes, and pathways.

## Results

### Genetic Correlation between Endometriosis and Ovarian Cancer Histotypes

Genetic correlation can be used to describe the genetic relationship between two traits and is an estimate of the proportion of variance that two traits share that is attributed to genetics. Estimating the genetic correlation between traits contributes to our understanding of shared underlying genetic risk factors and biological pathways. To estimate the genetic correlation between epithelial ovarian cancer (EOC) histotypes and endometriosis we used GWAS summary statistics from meta-analyses conducted by Phelan *et al*. (2017) and Sapkota *et al*. (2017) respectively, and linkage disequilibrium (LD) score regression (Bulik-Sullivan *et al*, 2015). SNPs were matched on position and alleles to ensure effect size estimates were harmonised across data sets to obtain a set of 7,617,581 SNPs represented in the EOC histotypes and endometriosis datasets. We estimated significant (P-value < 0.05) positive genetic correlations (r_g_) between endometriosis and CCOC (r_g_=0.71), ENOC (r_g_=0.48) and HGSOC (r_g_=0.19) (Table 1). The r_g_ for genetic correlation with LMPSOC was 0.88 but this did not reach statistical significance, and we were unable to estimate r_g_ for LGSOC due to this histotype having the smallest sample size (1,012 cases). There was no evidence of a significant correlation between MOC and endometriosis.

**Table 1.** Genetic correlation (r_g_) between endometriosis and epithelial ovarian cancer (EOC) histotypes.

| EOC Histotype | $r_g$ (se) | P-value |
| --- | --- | --- |
| Clear cell | 0.71 (0.26) | 0.007 |
| Endometrioid | 0.48 (0.20) | 0.016 |
| High-grade Serous | 0.19 (0.09) | 0.033 |
| Low-grade Serous | NA | NA |
| Low malignant potential serous | 0.88 (0.85) | 0.401 |
| Mucinous | -0.18 (0.15) | 0.227 |
| se = standard error |  |  |

### Mendelian Randomization Analysis

We then used Mendelian randomization based on the inverse-variance weighted (IVW) method (Burgess *et al*, 2013) and sensitivity analyses based on the weighted median (Bowden *et al*, 2016) and MR-Egger (Bowden *et al*, 2015) methods to investigate the association between genetic liability to endometriosis and EOC histotypes. Genetic liability to endometriosis as predicted by 25 independent genome-wide significant (Rahmioglu *et al*, 2018) (P < 5×10^−8^) endometriosis lead SNPs was associated with increased risk of CCOC, ENOC, HGSOC and LMPSOC in the IVW analysis and the results were robust in sensitivity analyses (Table 2). The strongest associations were observed for ENOC (P = 1.4×10^−10;^ OR=1.66 (1.42-1.93)) and CCOC (P = 2.8×10^−18^, OR=2.59 (2.09-3.21)).

**Table 2.** Mendelian randomization results considering genetic liability to endometriosis as the exposure and epithelial ovarian cancer (EOC) histotypes as the outcome.

| EOC Histotype | MR Method | OR (95% CI) | P-value |
| --- | --- | --- | --- |
| High Grade Serous | IVW | 1.22 (1.07-1.38) | 0.002 |
|  | Weighted Median | 1.16 (1.02-1.32) | 0.025 |
|  | MR-Egger | 1.23 (0.82-1.86) | 0.319 |
| Low Grade Serous | IVW | 1.27 (0.96-1.67) | 0.091 |
|  | Weighted Median | 1.27 (0.87-1.84) | 0.212 |
|  | MR-Egger | 0.86 (0.35-2.09) | 0.742 |
| Low Malignant Potential Serous | IVW | 1.45 (1.17-1.79) | 0.001 |
|  | Weighted Median | 1.52 (1.16-1.99) | 0.003 |
|  | MR-Egger | 1.42 (0.71-2.83) | 0.323 |
| Mucinous | IVW | 1.24 (1-1.53) | 0.046 |
|  | Weighted Median | 1.03 (0.77-1.39) | 0.821 |
|  | MR-Egger | 0.83 (0.42-1.63) | 0.583 |
| Endometrioid | IVW | 1.66 (1.42-1.93) | 1.4E-10 |
|  | Weighted Median | 1.58 (1.27-1.97) | 3.0E-05 |
|  | MR-Egger | 1.63 (1-2.67) | 0.051 |
| Clear Cell | IVW | 2.59 (2.09-3.21) | 2.8E-18 |
|  | Weighted Median | 2.48 (1.82-3.39) | 9.6E-08 |
|  | MR-Egger | 2.54 (1.28-5.02) | 0.007 |
| OR = Odds Ratio |  |  |  |
| 95% CI = 95% Confidence Interval |  |  |  |

### Genetic Associations shared between Endometriosis and Ovarian Cancer Histotypes

To identify genetic associations with some evidence of a shared contribution from both diseases, we combined the EOC histotypes and endometriosis susceptibility data sets using two complementary approaches; first, meta-analysis using approximate Bayes factors computed and combined by the MetABF method in both an independent and fixed model (Trochet *et al*, 2019) and second, meta-analysis based on the modified Han and Eskin random-effects model and fixed effects model implemented in RE2C (Han & Eskin, 2011; Lee *et al*, 2017). The cross-trait meta-analysis identified several genome-wide significant associations and a summary of the number of SNPs nominally associated with both endometriosis and each ovarian cancer histotype using MetABF and RE2C are listed in Table 3. SNPs were considered as markers of a shared genetic association with both traits if they had (i) a log_10_ ABF > 4 in the cross-trait MetABF analysis using either model, (ii) a P-value < 5×10^−8^ in the cross-trait RE2C analysis using either model, and (iii) a P-value < 0.05 in each single trait meta-analysis. A combined log_10_ ABF > 4 is equivalent to a posterior probability of combined association > 90% given a prior probability of association at any SNP of 1 in 1,000. All SNPs (n=2,237 non-redundant) with P-value < 5×10^−8^ in RE2C had log_10_ ABF > 4 in MetABF suggesting good consistency between the methods. Filtering out SNPs that did not have evidence for nominal association in each single trait meta-analysis (P < 0.05) filtered out ∼68% of the 3,612 SNPs, leaving 1,144 SNPs that met all three aforementioned criteria. The largest number of shared genome-wide significant loci (or regions) were identified between endometriosis and CCOC (14 loci; Tables 3 & 4). This was followed by 13 risk loci shared between endometriosis and HGSOC, six risk loci with ENOC, five risk loci with MOC, five risk loci with LMPSOC and three risk loci with LGSOC (Tables 3 & 4). Four loci had lead SNPs with opposite directions of allelic association between endometriosis and the EOC histotype (Table 4). Significant SNPs in each analysis are listed in Supplementary Tables 1 & 2. Several loci also contain lead SNPs that have been associated with other reproductive traits and diseases including uterine fibroids, sex hormone levels, polycystic ovarian syndrome (PCOS) and age at menarche (Supplementary Table 3).

**Table 3.**
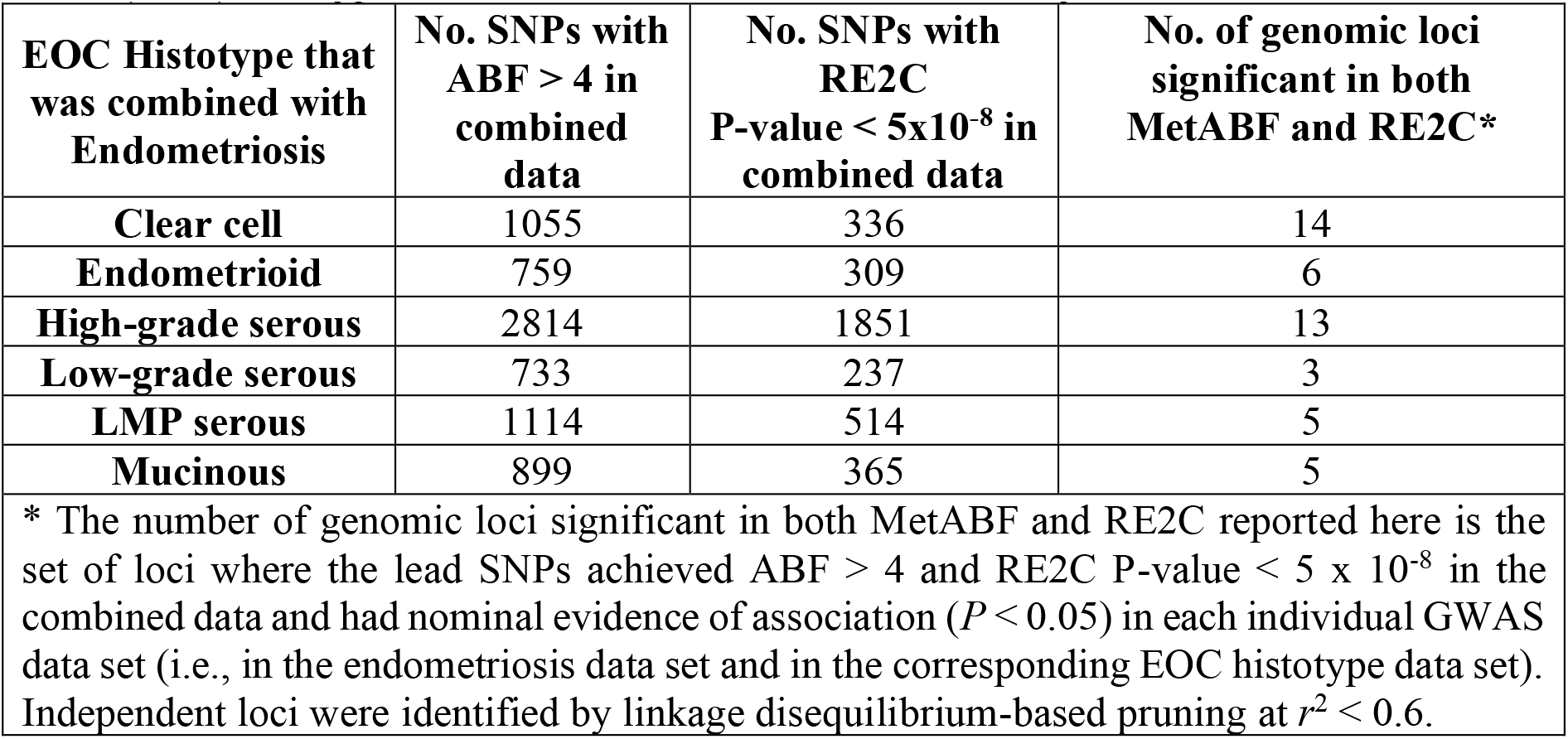
Number of significant SNPs and genomic loci identified in the epithelial ovarian cancer (EOC) histotype and endometriosis cross-trait meta-analyses.

| EOC Histotype that was combined with Endometriosis | No. SNPs with ABF > 4 in combined data | No. SNPs with RE2C P-value < 5x10<sup>-8</sup> in combined data | No. of genomic loci significant in both MetABF and RE2C* |
| --- | --- | --- | --- |
| **Clear cell** | 1055 | 336 | 14 |
| **Endometrioid** | 759 | 309 | 6 |
| **High-grade serous** | 2814 | 1851 | 13 |
| **Low-grade serous** | 733 | 237 | 3 |
| **LMP serous** | 1114 | 514 | 5 |
| **Mucinous** | 899 | 365 | 5 |

\* The number of genomic loci significant in both MetABF and RE2C reported here is the set of loci where the lead SNPs achieved ABF > 4 and RE2C P-value < 5 x 10<sup>-8</sup> in the combined data and had nominal evidence of association ( $P < 0.05$ ) in each individual GWAS data set (i.e., in the endometriosis data set and in the corresponding EOC histotype data set). Independent loci were identified by linkage disequilibrium-based pruning at $r^2 < 0.6$ .

**Table 4.**
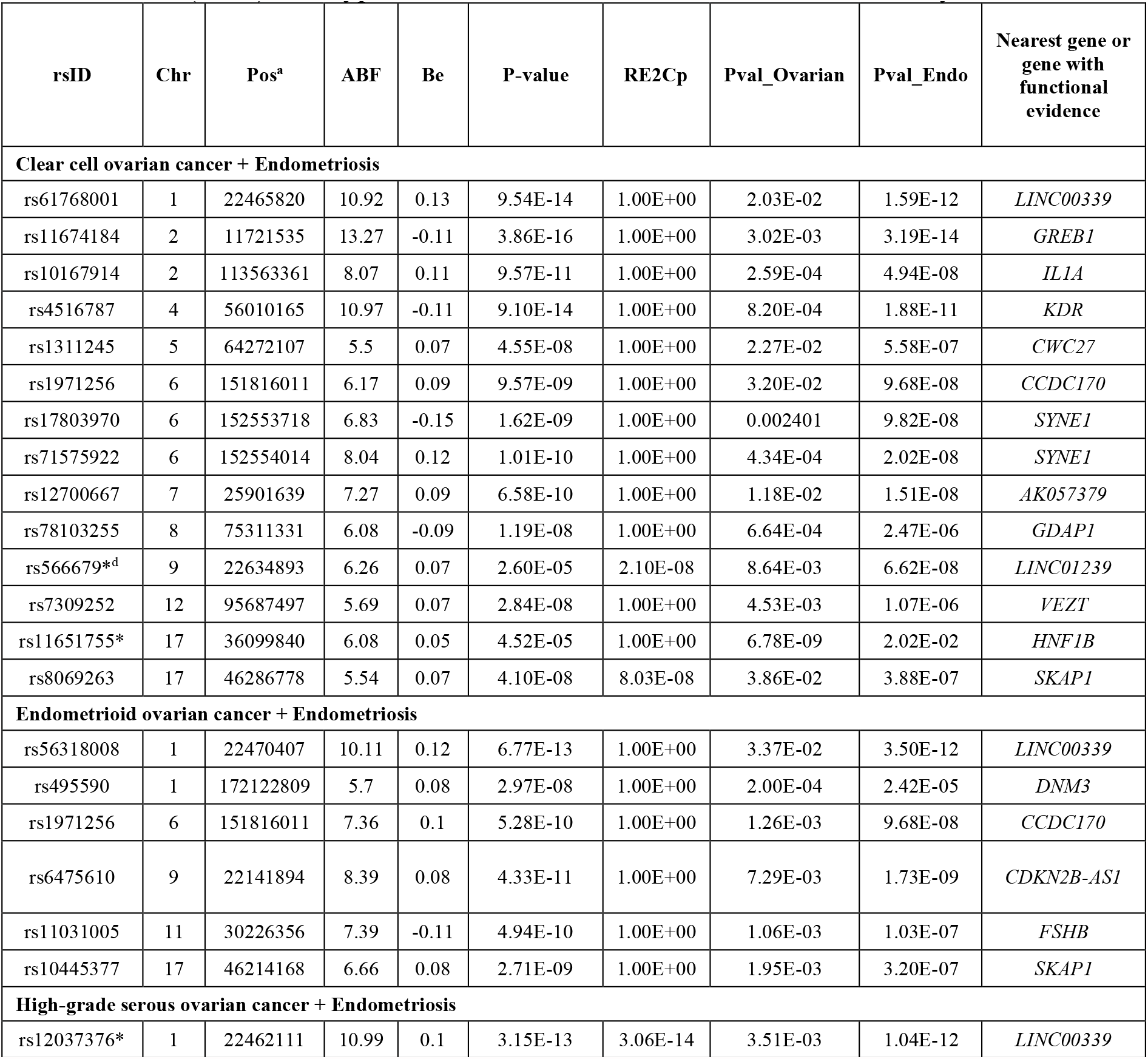
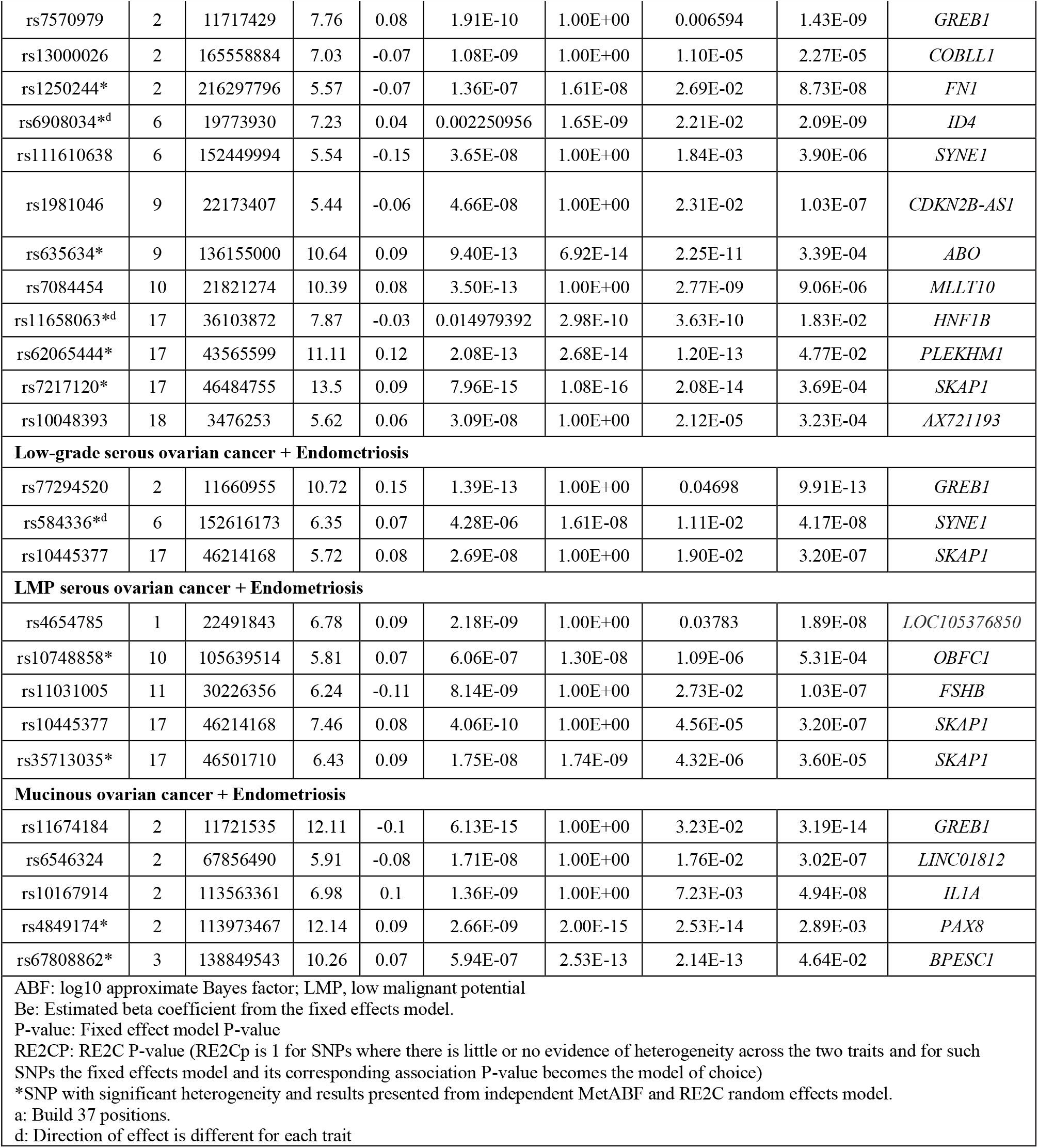
Lead SNPs in genomic loci that demonstrated shared associations with an epithelial ovarian cancer (EOC) histotype and endometriosis from the RE2C meta-analysis.

### Colocalization Analyses to identify Shared Causal Variants

Our MetABF and RE2C analyses identified shared susceptibility loci for endometriosis and EOC. However, it is not clear whether the same candidate causal variants underlie the associations at these loci or whether the associations at these loci are driven by distinct candidate causal variants for endometriosis and EOC. We examined the underlying shared genetic architecture of endometriosis and EOC further using a statistical model to estimate the posterior probability of association (PPA) that a genomic region 1) contains a variant associated only with endometriosis (PPA_1), 2) contains a variant associated only with ovarian cancer (PPA_2), 3) contains a variant associated with both traits (PPA_3) and 4) contains both a variant associated with endometriosis and an independent variant associated with ovarian cancer (PPA_4). These models were implemented in gwas-pw (Pickrell *et al*, 2016). Genomic regions with a PPA_3>0.5, evidence of a candidate causal variant influencing both diseases, or PPA_4>0.5, evidence that the candidate causal variants underlying the association with each trait were distinct, are listed in Table 5. CCOC had the largest number of genomic regions (n=13) with evidence of a shared causal variant with endometriosis. Figure 1 shows the genomic regions where there was statistical evidence for the same causal variant underpinning endometriosis and at least one EOC histotype or where there was evidence for two distinct signals. All regions identified with PPA_3 or PPA_4 >0.5 contained lead SNPs significant in the cross-trait meta-analyses (log_10_ ABF > 4 in the cross-trait MetABF analysis, P-value < 5×10^−8^ in the cross-trait RE2C analysis and P-value < 0.05 in each single trait meta-analysis) except for one region on chromosome 3 (Chr 3:126215130-128194265) where only colocalization offered evidence for a shared association between MOC and endometriosis. Two regions with PPA_3>0.5 that achieved genome-wide significance in the meta-analyses (P-value < 5×10^−8^ and log_10_ABF > 4) were > 1 Mb from any risk locus previously reported for endometriosis and EOC: 2q24.3 (rs13000026) and 18p11.31 (rs10048393). One of the four loci (9p21) with lead SNPs with opposite directions of effect, identified in the cross-trait meta-analysis between endometriosis and CCOC, also had evidence for the same causal variant underpinning both diseases from the colocalization analysis. Another on chromosome 17 (17q12) had evidence for two distinct signals for endometriosis and HGSOC. The remaining two had no evidence of colocalization. Several genomic regions containing genome-wide significant associations identified in the cross-trait meta-analyses only achieved PPA_1/2>0.5 suggesting the associations were only driven by one of the two traits. However, this can also occur due to the limited power to detect colocalization with the smaller sample sizes that were available for cross-trait colocalization analyses involving the less common EOC histotypes.

**Table 5.**
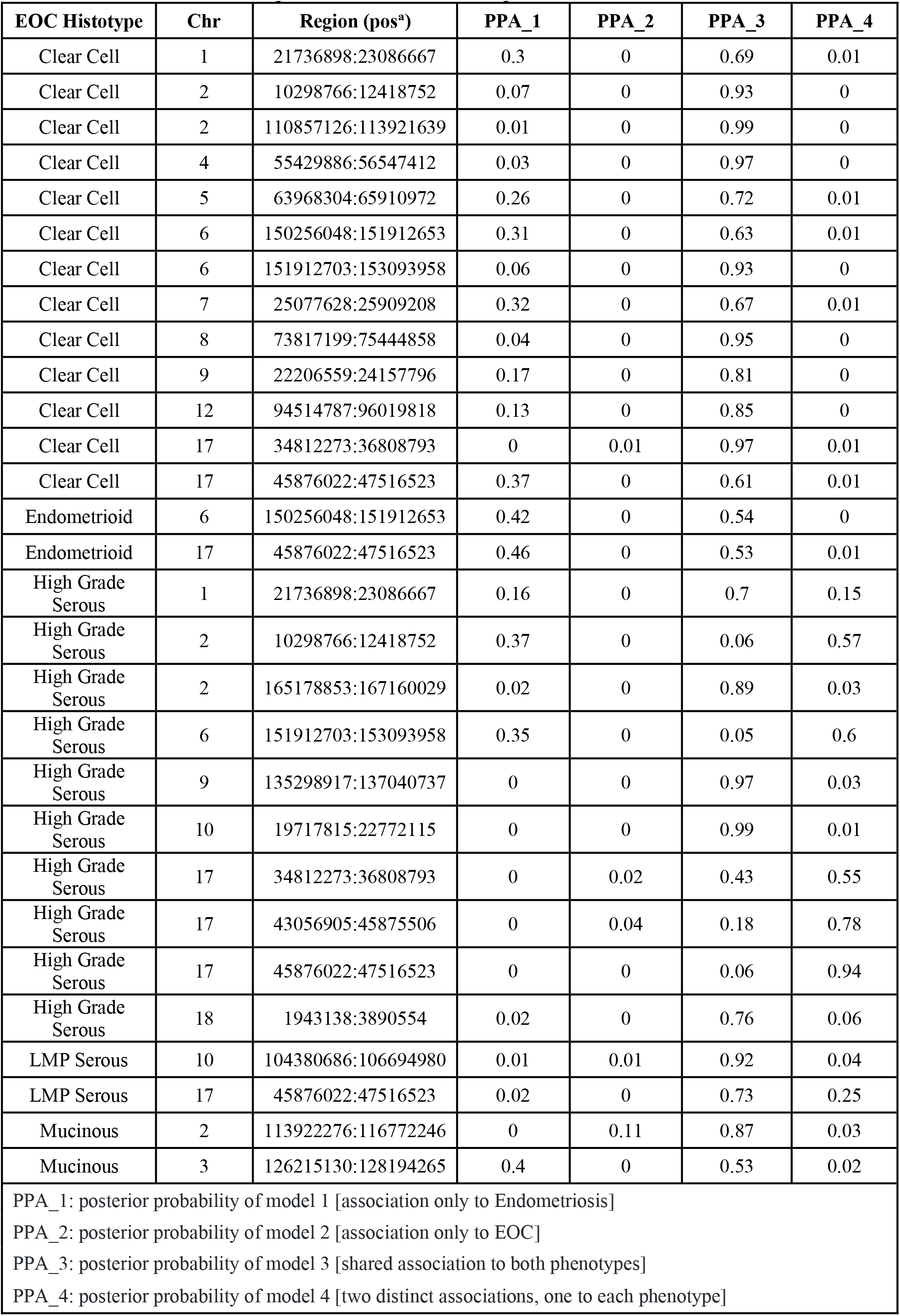
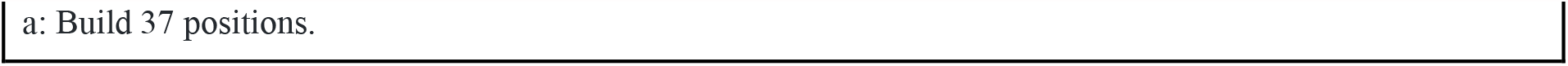
GWAS-pw results for analyses between epithelial ovarian cancer (EOC) histotypes and endometriosis. Posterior probabilities of GWAS-pw models.

| EOC Histotype | Chr | Region (pos<sup>a</sup>) | PPA_1 | PPA_2 | PPA_3 | PPA_4 |
| --- | --- | --- | --- | --- | --- | --- |
| Clear Cell | 1 | 21736898:23086667 | 0.3 | 0 | 0.69 | 0.01 |
| Clear Cell | 2 | 10298766:12418752 | 0.07 | 0 | 0.93 | 0 |
| Clear Cell | 2 | 110857126:113921639 | 0.01 | 0 | 0.99 | 0 |
| Clear Cell | 4 | 55429886:56547412 | 0.03 | 0 | 0.97 | 0 |
| Clear Cell | 5 | 63968304:65910972 | 0.26 | 0 | 0.72 | 0.01 |
| Clear Cell | 6 | 150256048:151912653 | 0.31 | 0 | 0.63 | 0.01 |
| Clear Cell | 6 | 151912703:153093958 | 0.06 | 0 | 0.93 | 0 |
| Clear Cell | 7 | 25077628:25909208 | 0.32 | 0 | 0.67 | 0.01 |
| Clear Cell | 8 | 73817199:75444858 | 0.04 | 0 | 0.95 | 0 |
| Clear Cell | 9 | 22206559:24157796 | 0.17 | 0 | 0.81 | 0 |
| Clear Cell | 12 | 94514787:96019818 | 0.13 | 0 | 0.85 | 0 |
| Clear Cell | 17 | 34812273:36808793 | 0 | 0.01 | 0.97 | 0.01 |
| Clear Cell | 17 | 45876022:47516523 | 0.37 | 0 | 0.61 | 0.01 |
| Endometrioid | 6 | 150256048:151912653 | 0.42 | 0 | 0.54 | 0 |
| Endometrioid | 17 | 45876022:47516523 | 0.46 | 0 | 0.53 | 0.01 |
| High Grade Serous | 1 | 21736898:23086667 | 0.16 | 0 | 0.7 | 0.15 |
| High Grade Serous | 2 | 10298766:12418752 | 0.37 | 0 | 0.06 | 0.57 |
| High Grade Serous | 2 | 165178853:167160029 | 0.02 | 0 | 0.89 | 0.03 |
| High Grade Serous | 6 | 151912703:153093958 | 0.35 | 0 | 0.05 | 0.6 |
| High Grade Serous | 9 | 135298917:137040737 | 0 | 0 | 0.97 | 0.03 |
| High Grade Serous | 10 | 19717815:22772115 | 0 | 0 | 0.99 | 0.01 |
| High Grade Serous | 17 | 34812273:36808793 | 0 | 0.02 | 0.43 | 0.55 |
| High Grade Serous | 17 | 43056905:45875506 | 0 | 0.04 | 0.18 | 0.78 |
| High Grade Serous | 17 | 45876022:47516523 | 0 | 0 | 0.06 | 0.94 |
| High Grade Serous | 18 | 1943138:3890554 | 0.02 | 0 | 0.76 | 0.06 |
| LMP Serous | 10 | 104380686:106694980 | 0.01 | 0.01 | 0.92 | 0.04 |
| LMP Serous | 17 | 45876022:47516523 | 0.02 | 0 | 0.73 | 0.25 |
| Mucinous | 2 | 113922276:116772246 | 0 | 0.11 | 0.87 | 0.03 |
| Mucinous | 3 | 126215130:128194265 | 0.4 | 0 | 0.53 | 0.02 |
| PPA_1: posterior probability of model 1 [association only to Endometriosis] |  |  |  |  |  |  |
| PPA_2: posterior probability of model 2 [association only to EOC] |  |  |  |  |  |  |
| PPA_3: posterior probability of model 3 [shared association to both phenotypes] |  |  |  |  |  |  |
| PPA_4: posterior probability of model 4 [two distinct associations, one to each phenotype] |  |  |  |  |  |  |

a: Build 37 positions.

**Figure 1.**
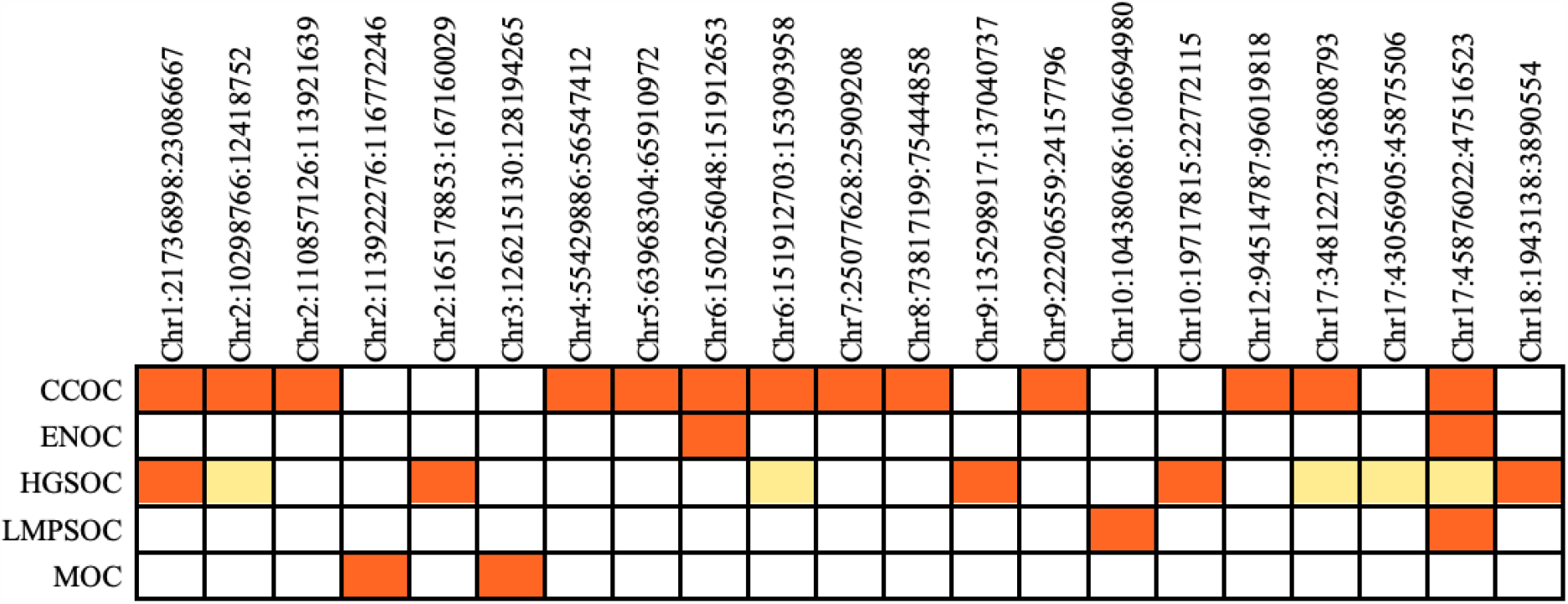
Genomic regions associated with risk of endometriosis and epithelial ovarian cancer (EOC) histotypes. Regions marked as orange have a posterior probability of model 3 (PPA_3) > 0.5 evidence that the same underlying candidate causal variant is associated with endometriosis and the ovarian cancer histotype in that region. Regions marked as yellow have a posterior probability of model 4 (PPA_4) > 0.5 evidence that the region contains a variant associated with endometriosis and an independent variant associated with ovarian cancer histotype. All regions shaded in orange contained genome-wide significant loci in the cross-trait meta-analysis except Chr 3:126215130-128194265.

### Gene-based Association Analysis of Endometriosis and Ovarian Cancer Histotypes

We conducted a gene-based association analysis using fastBAT (Bakshi *et al*, 2016), a statistical association test that calculates the combined association for all SNPs mapped to each gene while taking into account correlation between SNPs due to LD. Nine genes were associated at genome-wide significance (P-value < 2.45×10^−6^) with endometriosis (*GREB1, MIR4429, KDR, WNT4, SYNE1, CDKN2B-AS1, CDC42, ID4, PTPRO*), 67 with HGSOC, one with LGSOC (*KIAA1024*), four for LMPSOC (*TERT, SLC6A18, MIR4457, CLPTM1L*) and 27 for MOC in single trait gene-based analysis (Supplementary Table 4). Genome-wide significant genes for endometriosis were nominally significant (P < 0.05) for CCOC (*GREB1, MIR4429, WNT4*), ENOC (*CDNK2B-AS1*) and HGSOC (*CDNK2B-AS1, MIR4429, WNT4*) (Supplementary Table 4).

We looked at the overlap between the top 1% of genes associated with each trait (204/20,439 genes evaluated in the fastBAT analysis) and observed an overlap of 5% between endometriosis and HGSOC (11 genes), 4% with CCOC (9 genes), 3% with LMPSOC (7 genes), 3% with ENOC (6 genes), 3% with MOC (5 genes) and 1% with LGSOC (2 genes). Two genes, *SNX11* and *CBX1*, were associated with endometriosis, ENOC, HGSOC and LMPSOC. *SKAP1* was associated with HGSOC, LMPSOC and endometriosis. However, none of the genes in the top 1% that overlapped between endometriosis and CCOC were in the top 1% genes associated with other histotypes. Using an over-representation analysis in WebGestalt (Liao *et al*, 2019), no specific pathways were significantly enriched (FDR < 0.05) for overlapping genes. This was also the case when the analysis was extended to the top 5% of genes associated with each trait and the overlapping genes between endometriosis and each ovarian cancer histotype in the top 5% considered (Supplementary Table 4).

### Functional Annotation

We collated all candidate causal variants by identifying all SNPs in tight linkage disequilibrium (LD) with the lead SNPs (r^2^>0.7) from the cross-trait meta-analyses (log_10_ ABF > 4 in the cross-trait MetABF analysis, P-value < 5×10^−8^ in the cross-trait RE2C analysis and P-value < 0.05 in each single trait meta-analysis). The set of candidate causal variants included 4,044 unique SNPs, which we functionally annotated to genes and epigenomic biofeatures:

#### Overlap with noncoding DNA biofeatures

To identify putative functional SNPs, we overlapped all candidate causal SNPs with noncoding regulatory elements (biofeatures) identified by epigenomic profiling of disease-relevant tissues and cell lines. The biofeature catalogue consists of 11 consensus peak sets (see Methods, Supplementary Table 5) derived from 45 epigenomic profiles. Epigenome features included open chromatin (18 ATAC-seq data sets) and active chromatin (27 H3K27ac ChIP-seq profiles; Supplementary Table 6). The specimens profiled include non-cancerous gynecologic tissues (fallopian tube, endometriosis and endometriosis-associated stroma) and ovarian cancer (clear cell, endometrioid, high-grade serous and mucinous) tissues or cell line models (Coetzee *et al*, 2015b; Corona *et al*, 2020a). Consensus peak sets averaged 33.6 (sd = 22, range = [9,84.3]) thousand peaks spanning, on average, 1.04% of the human genome (sd = 0.37, range = [0.42, 1.53]) (Supplementary Figure 1a-c, Supplementary Table 6). Genome coverage is marginally correlated with number of donors (Spearman’s rho = 0.43, p-value = 0.18; Supplementary Figure 1d-f).

We reduced the 1,144 candidate SNPs to 824 non-redundant variants most strongly associated with both endometriosis and EOC histotypes (log10 ABF > 4 in the cross-trait MetABF analysis, a P-value < 5×10^−8^ in the cross-trait RE2C analysis and a P-value < 0.05 in each individual trait meta-analysis). Of these 824 candidate causal variants, 119 (14.4%) overlapped at least one biofeature (Figure 2a, Supplementary Table 7). The proportion of independent loci containing SNPs intersecting with biofeatures varied by EOC histotype, with only 33.3% of loci associated with endometriosis plus LGSOC overlapping with at least one biofeature, while 71.4% of endometriosis and CCOC loci overlap with one or more relevant biofeatures (Table 8, Figure 2b, Supplementary Figure 2). As expected, ATAC-seq consensus peak sets provide different information compared to H3K27ac ChIP-seq peak sets. We observed that H3K27ac ChIP-seq consensus peak sets for fallopian tube, endometriosis-associated stroma and endometriosis primary tissues and ATAC-seq consensus peaks for CCOC and fallopian tube intersect a similar set of SNPs, possibly reflecting the epidemiologic links between these tissues and diseases (Supplementary Figure 3).

**Figure 2.**
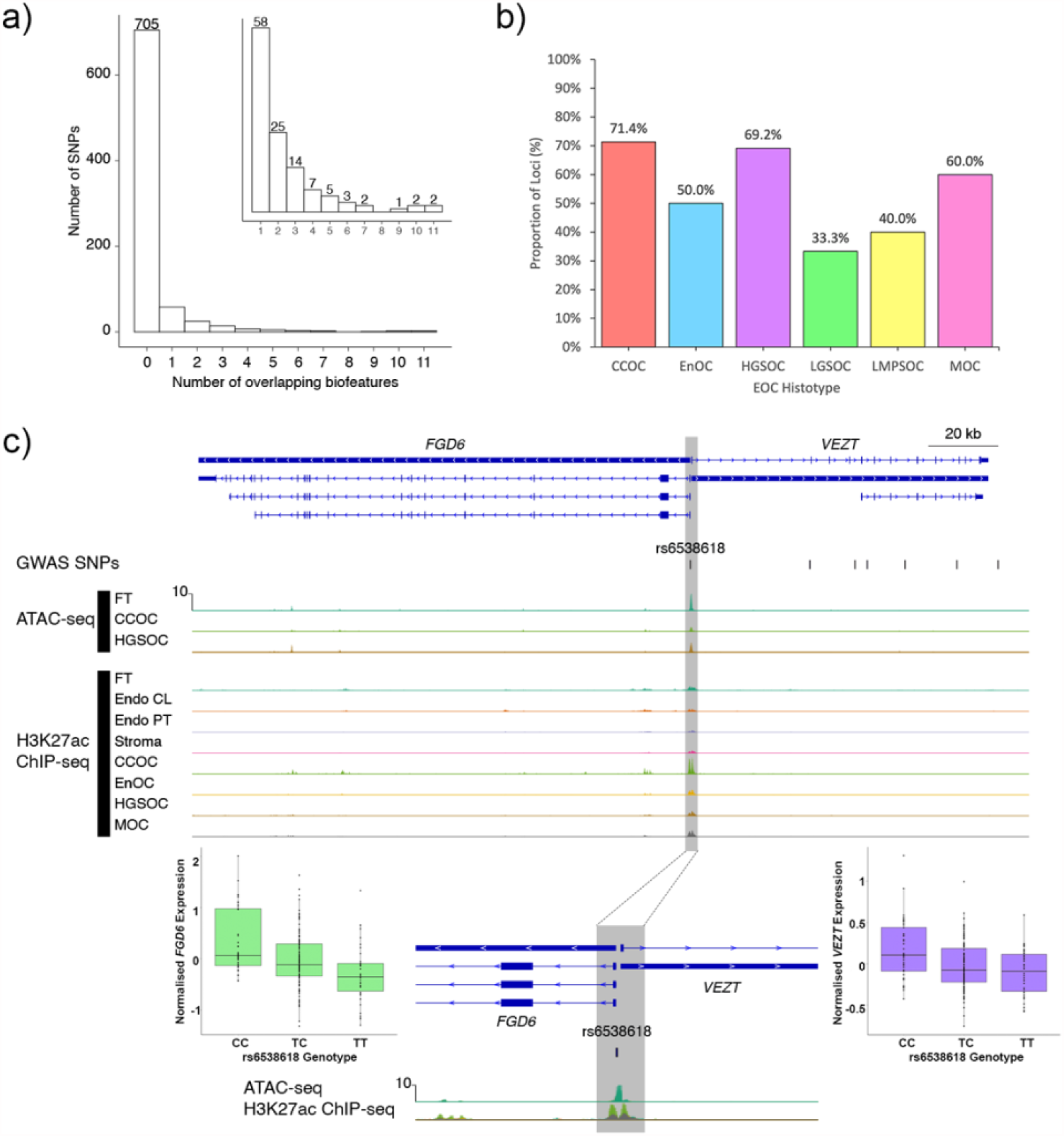
Functional annotation of SNPs associated with risk of endometriosis and ovarian cancer. (a) Histogram of number of non-redundant SNPs for all histological subtypes that overlap *n* biofeatures. Inset panel, histogram of number of non-redundant SNPs for all histological subtypes that overlap *≥* 1 biofeature. (b) Proportion of loci shared between endometriosis and each EOC histotype containing SNPs that overlap at least one biofeature. (c) A promoter SNP at the *VEZT/FGD6* locus overlaps 10 biofeatures and intersects with an active open region of chromatin that lies in a bidirectional promoter associated with these two genes. Biofeatures are shown as peaks on the ATAC-seq, H3K27ac and ChIP-seq tracks for primary tissues (PT) and cell lines (CL). Endo, endometriosis. Specimens are primary tissues unless otherwise indicated. The grey shaded area highlights peaks overlapping rs6538618. Boxplots show the association between rs6538618 genotypes and expression of *VEZT* and *FGD6* in endometrium.

The 119 SNPs that overlap at least one ‘consensus peak set’ are distributed across 28 distinct loci (Supplementary Table 7). Overlaps provide functional evidence that these SNPs in risk loci shared between endometriosis and EOC histotypes are located within regulatory regions. The *MLLT10* and *FSHB* loci contains the SNPs with the most functional evidence and highest number of overlaps, rs4071559 and rs10828247 each overlap eleven biofeatures (Supplementary Table 7). The *VEZT* locus harbors the SNPs with the second highest number of overlaps, where rs6538618 overlaps ten biofeatures at the putative bidirectional *VEZT/FGD6* promoter (Figure 2c). rs6538618 has additional functional evidence and has been associated with the expression of both *VEZT* and *FGD6* in endometrium (Mortlock *et al*, 2020b), fibroblasts, artery and muscle tissue (Consortium *et al*, 2017) (Figure 2c). The *SKAP1* and *PAX8* contain the greatest number of SNPs overlapping biofeatures (26 SNPs).

#### Tissue specific effects and disease relevant pathways

Using Functional Mapping and Annotation (FUMA)(Watanabe *et al*, 2017) we identified that the expression of genes containing, or nearby to, SNPs shared between endometriosis and two ovarian cancer histotypes (CCOC & HGSOC) clustered across reproductive tissues including ovary, fallopian tube and uterus (Supplementary Figure 4). Several pathways were enriched within the set of genes annotated to significant SNPs (Supplementary Table 9). Unlike the fastBAT analysis genes were not identified using a gene-based association analysis (SNPs within gene) but were instead annotated based on position (gene within 10kb of SNP). Focusing on enriched pathways containing three or more genes, pathways related to cell adhesion and nuclear division were enriched for genes annotated to SNPs associated with both endometriosis and CCOC. Gene sets associated with other reproductive traits and diseases were also enriched including uterine fibroids, endometrial cancer, dysmenorrheic pain severity and gestational age at birth (Supplementary Table 9).

#### Causal associations with gene expression and methylation

The fastBAT analysis involved a purely statistical gene-level association test. To complement fastBAT, we used Summary-data-based Mendelian Randomization (SMR)(Zhu *et al*, 2016) that integrates gene-level expression and methylation with the GWAS data to elucidate potential gene-level functional mechanisms. SMR enables the identification of potentially causal associations between shared susceptibility to endometriosis and ovarian cancer histotypes and gene expression using SNPs associated with the traits from their individual GWA meta-analyses. We performed an SMR analysis using summary statistics from the endometriosis GWAS meta-analysis, each of the ovarian cancer histotype and eQTL data from endometrium (Fung *et al*, 2017; Mortlock *et al*, 2020a), blood (Võsa *et al*, 2018) and GTEx uterus and ovary (Consortium *et al*., 2017) (Supplementary Table 10). Associations (SMR P-value < 0.05; Heterogeneity In Dependent Instruments (HEIDI) P-value > 0.05) were identified between variants, risk of endometriosis and at least one EOC histotype and expression (P-value < 5×10^−8^) of 18 genes in endometrium, 25 genes in the ovary and 10 genes in the uterus, the majority of genes only shared with a single histotype. This was not dependent on the same variant effecting the gene in both diseases. Variants were associated with risk of endometriosis, CCOC, ENOC and expression of *HLA-C* in both the ovary and endometrium and risk of endometriosis and HGSOC and expression of *HLA-K* in ovary and uterus.

Previous studies have shown a large proportion of eQTLs are shared between tissues (Consortium *et al*., 2017; Mortlock *et al*., 2020b). To increase power the analysis was repeated using a large blood *cis*-eQTL dataset from eQTLGen (Võsa *et al*., 2018) (n=31,684 individuals) as a proxy and expression of 244 genes were associated (SMR P-value < 0.05; HEIDI P-value > 0.05) with both risk of endometriosis and ovarian cancer histotypes (Supplementary Table 10). Some of the top associations based on the blood eQTL data include expression of *SKAP1* and risk of endometriosis, LMPSOC, CCOC and LGSOC, expression of *TNPO3* and *IRF5* and risk of endometriosis, HGSOC and LMPSOC and expression of *CEP97* and risk of endometriosis and CCOC.

Significant associations (SMR P-value < 0.05; HEIDI P-value > 0.05) between variants, methylation in the endometrium, and risk of endometriosis and at least one EOC histotype were identified at 112 CpG sites including those near the *GREB1* signal for endometriosis, CCOC, ENOC, HGSOC and MOC (Supplementary Table 10). Using a large blood methylation QTL dataset (McRae *et al*, 2018) for SMR analysis, we identified variants affecting methylation at 1,449 CpG sites where variants associated with methylation were also associated with endometriosis and at least one EOC histotype including sites near *ESR1* for CCOC, ENOC and MOC, *SYNE1* for ENOC, HGSOC and LMPSOC, *SKAP1* for CCOC, ENOC and LGSOC and *MLLT10* for CCOC, HGSOC, LGSOC and LMPSOC. Table 7 summarises the various levels of evidence gained from the aforementioned analyses for loci associated with both endometriosis and EOC histotypes.

**Table 6.**
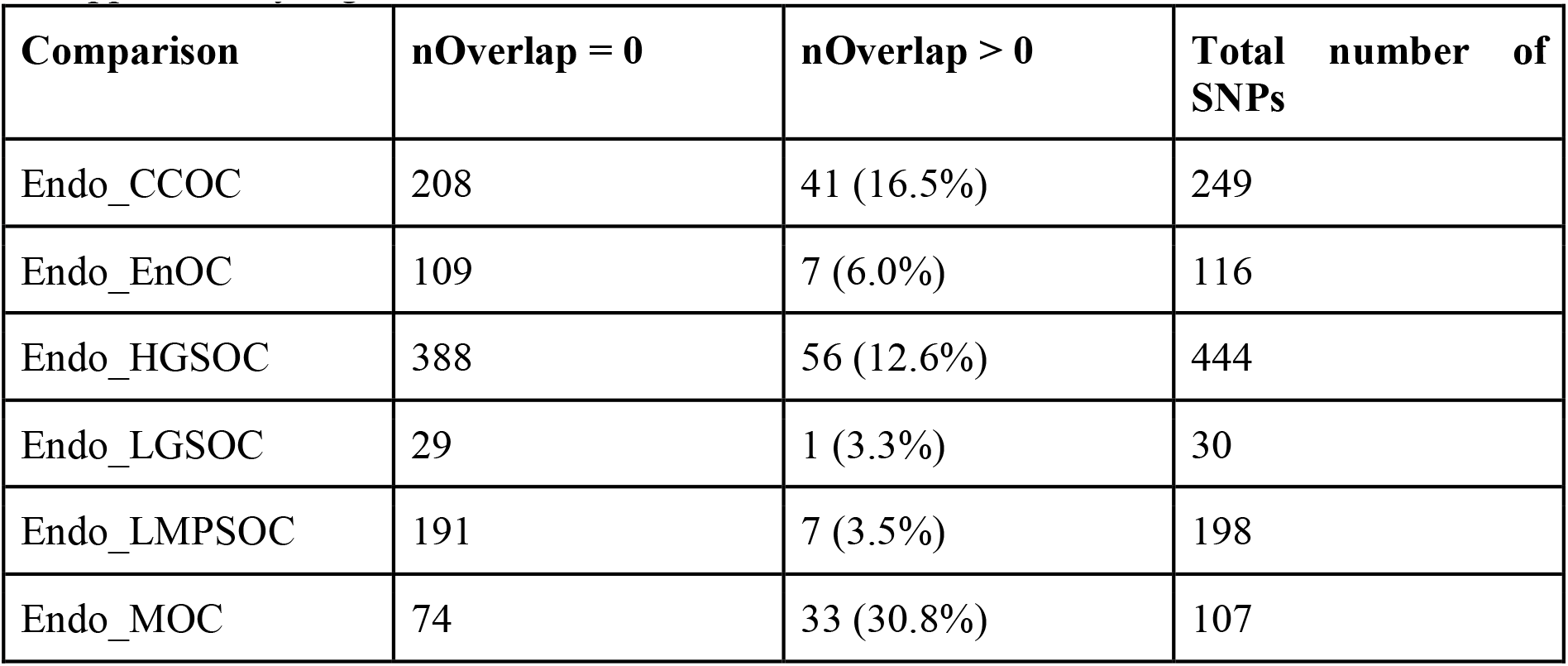
Number of SNPs overlapping zero or more than zero consensus peak sets. Related to Supplementary Figure 4.

| Comparison | nOverlap = 0 | nOverlap > 0 | Total number of SNPs |
| --- | --- | --- | --- |
| Endo_CCOC | 208 | 41 (16.5%) | 249 |
| Endo_EnOC | 109 | 7 (6.0%) | 116 |
| Endo_HGSOC | 388 | 56 (12.6%) | 444 |
| Endo_LGSOC | 29 | 1 (3.3%) | 30 |
| Endo_LMPSOC | 191 | 7 (3.5%) | 198 |
| Endo_MOC | 74 | 33 (30.8%) | 107 |

**Table 7.** Summary of evidence from the bivariate meta-analysis, GWAS-pw, overlap of biofeatures, fastBAT and Summary-data-based Mendelian Randomisation (SMR) for loci associated with both endometriosis and epithelial ovarian cancer (EOC) histotypes. Cells shaded orange contain EOC histotypes with a posterior probability of a shared variant association with endometriosis of >0.5, those shaded yellow contain EOC histotypes with a posterior probability of two distinct associations, one to each phenotype, of >0.5. The blue colour gradient in the feature overlap column corresponds to the maximum number of feature overlaps identified for a variant in that locus.

| Risk SNP (cytoband) | Significant in Meta-analysis | Colocalised Signal | Feature Overlap | fastBAT | SMR |
| --- | --- | --- | --- | --- | --- |
| rs495590 (1q24.3) | ENOC |  |  | *DNM3* |  |
| rs61768001/rs12037376 (1p36.12) | CCOC, HGSOC | CCOC, HGSOC | 5 | *WNT4* | CpG site near *WNT4* |
| rs10167914 (2q13) | CCOC, MOC | CCOC | 3 | *IL1A* |  |
| rs4849174 (2q13) | MOC | MOC | 7 | *PSD4* |  |
| rs13000026 (2q24.3) | HGSOC | HGSOC |  |  |  |
| rs11674184 (2p25.1) | CCOC, MOC | CCOC | 2 | *GREB1/MIR4429* |  |
| rs7570979 (2p25.1) | HGSOC | HGSOC | 2 | *MIR4429* |  |
| rs1250244 (2q35) | HGSOC |  | 2 |  |  |
| rs4516787 (4q12) | CCOC | CCOC | 1 |  |  |
| rs1311245 (5q12.3) | CCOC | CCOC | 1 |  |  |
| rs6908034 (6p22.3) | HGSOC |  |  |  |  |
| rs1971256 (6q25.1) | CCOC, ENOC | CCOC, ENOC | 5 | *CCDC170* |  |
| rs17803970 (6q25.2) | CCOC | CCOC |  |  |  |
| rs71575922 (6q25.2) | CCOC | CCOC |  |  |  |
| rs111610638 (6q25.2) | HGSOC | HGSOC | 1 |  | CpG site near *SYNE1* |
| rs12700667 (7p15.2) | CCOC | CCOC | 2 |  |  |
| rs78103255 (8q21.11) | CCOC | CCOC |  | *GDAP1* | *GDAP1* expression<br>CpG site near *GDAP1* |
| rs566679 (9p21.3) | CCOC | CCOC |  |  |  |
| rs6475610 (9p21.3) | ENOC |  |  | *CDKN2B-AS1/CDKN2A* |  |
| rs1981046 (9p21.3) | HGSOC |  |  | *CDKN2B-AS1* |  |
| rs635634 (9q34.2) | HGSOC | HGSOC | 4 |  | *ABO* expression |
| rs7084454 (10p12.31) | HGSOC | HGSOC | 11 | *MLLT10/CASC10/SKIDA1/DNAJC1* | CpG site near *MLLT10* |
| rs10748858 (10q24.33) | LMPSOC | LMPSOC |  | *OBFC1* |  |
| rs11031005 (11p14.1) | ENOC, LMPSOC |  | 11 |  | CpG site near *FSHB*<br>*ARL14EP* expression |
| rs7309252 (12q22) | CCOC | CCOC | 10 | *VEZT* |  |
| rs11651755 (17q12) | CCOC | CCOC | 3 |  | CpG site near *HNF1B* |
| rs11658063 (17q12) | HGSOC | HGSOC | 3 |  | CpG site near *HNF1B* |
| rs62065444 (17q21.31) | HGSOC | HGSOC |  |  | CpG site near *ARHGAP27* |
| rs8069263/rs10445377 (17q21.32) | CCOC, ENOC, LGSOC, LMPSOC | CCOC, ENOC, LMPSOC | 2 | *SKAP1/SNX11/CBX1/NFE2L1/LOC101927166* | CpG site near *HOXB8*<br>CpG site near *SKAP1* |
| rs7217120 (17q21.32) | HGSOC | HGSOC | 10 | *SKAP1/SNX11/CBX1/NFE2L1/LOC101927166/HOXB2* |  |
| rs35713035 (17q21.32) | LMPSOC | LMPSOC | 3 | *SKAP1/SNX11/CBX1/NFE2L1/LOC101927166* | *SKAP1* expression |
| rs10048393 (18p11.31) | HGSOC | HGSOC | 1 | *LOC100505592* |  |

## Discussion

Analysis of germline genetic risk association data from endometriosis and ovarian cancer GWAS meta-analyses provides evidence of a genetic correlation and causal relationship between endometriosis and CCOC, ENOC and, to a lesser extent, HGSOC. Our results support epidemiological observations of an association between endometriosis and ovarian cancer as shown by estimates that women with endometriosis have two to three times higher risk of developing ovarian cancer (Pearce *et al*., 2012; Wei *et al*, 2011) and that a high proportion of CCOC and ENOC cases also have endometriosis (20-50%)(Jimbo *et al*., 1997; Stamp *et al*., 2016; Vercellini *et al*., 1993). Historically epidemiological studies have found little evidence for an association between endometriosis and HGSOC. However, a 2015 analysis that involved evaluating genetic loci known at the time to be associated with endometriosis risk in a smaller subset of the Ovarian Cancer Association Consortium case-control set used here found significant evidence using a gene-based statistical test of an association with both endometriosis and HGSOC risk at the 1p36 (*WNT4*) locus (Lee *et al*, 2016). Our Mendelian randomization (MR) results are consistent with findings from Yarmolinsky *et al* (2019). However, potentially due to our use of a larger number of SNPs to instrument endometriosis (25 SNPs based on the latest endometriosis GWAS versus 10 SNPs in the previously published analysis), we find that point estimates of the effect size for all associations in our analysis are larger than those reported in Yarmolinsky, et al. This is most notable in the odds ratio for CCOC (2.6 versus 1.5). The associations presented here reinforce the well-established links between endometriosis and endometrioid and clear cell EOCs and we also observe an association between endometriosis and HGSOC that was maintained across more than one analytic approach, suggesting some shared pathways underlie the development of these two phenotypes.

Using cross-trait meta-analyses we identify 28 distinct genomic loci that shared a lead variant contributing to the risk of both endometriosis and ovarian cancer histotypes. Colocalization analyses provided evidence (PPA_3>0.5) for a single causal variant underlying risk for both endometriosis and ovarian cancer in 19 of these regions. Functional annotation revealed that 14 of these 19 loci also contained risk SNPs that overlapped active and/or open chromatin. The high posterior probability of colocalization at a large number of distinct loci is a remarkable feature of the genetic relationship between endometriosis and EOC histotypes and suggests that identifying target genes in these loci may be valuable to understand the link between endometriosis and ovarian cancer and to intervene in neoplastic transformation.

Combining GWAS data for conditions known to predispose to cancers and for the corresponding cancers themselves has previously helped identify novel susceptibility loci for nevus density and melanoma (Duffy *et al*, 2018) and for gastroesophageal reflux disease and esophageal cancer (An *et al*, 2019). Our analysis identified two risk loci that are completely novel both in the context of endometriosis and EOC histotypes (i.e., > 1 Mb away from any previously identified locus). At the first locus, located at chromosome2q24.3, the index SNP (rs13000026) lies intronic to *Cordon-bleu protein-like 1* (*COBLL1*) and at the second locus at 18p11.31, index SNP rs10048393 lies intronic to a long noncoding RNA, *GAPLINC*. Whether these genes prove to be the target genes of these associations has yet to be determined; neither gene has been implicated in endometriosis nor ovarian cancer to date. The lead SNPs in these regions displayed strong associations (P-value <= 3.2×10^−4^) with endometriosis risk and HGSOC risk in the single trait GWAS data sets and the combined signal achieved genome-wide significance (P-value < 5×10^−8^). Moreover, gwas-pw colocalization analysis of each these new loci indicated a high probability (>= 0.76) of a single causal signal underlying the association with both traits.

Different regions shared between endometriosis and different histotypes may suggest possible biological mechanisms driving these causal relationships and the pathways contributing to risk of specific subtypes. Three regions identified as associated with both CCOC and endometriosis using bivariate meta-analysis and gwas-pw, chromosome 4 near *KDR*, chromosome 8 near *GDAP1* and chromosome 12 near *VEZT*, were not identified for ENOC and HGSOC. Similarly, genomic regions on chromosome 1 near *DNM3* and chromosome 11 near *FSHB* were associated with ENOC not CCOC or HGSOC, loci on chromosome 9 near *ABO* and chromosome 10 near *MLLT10* were associated with HGSOC not CCOC or ENOC. Alternatively, risk variants in the *SKAP1* locus on chromosome 17 were common between endometriosis and most histotypes. Shared variants in regions of known hormone responsive genes, oestrogen-responsive growth regulation by oestrogen in breast cancer 1 (*GREB1*)(Mohammed *et al*, 2013) and kinase insert domain receptor (*KDR*)(Sugino *et al*, 2002), may suggest a role of hormone regulation in the causal pathway between endometriosis and CCOC. Cell adhesion pathways were also significantly enriched for genes annotated to SNPs associated with risk of endometriosis and CCOC suggesting that the ability of cells to adhere may contribute to the pathogenesis of endometriosis and subsequently CCOC. Association between variants in the risk loci shared between endometriosis and EOC histotypes and other reproductive traits and diseases including PCOS, uterine fibroids and sex hormone levels suggests that perturbation of underlying pathways important for the development and regulation of the reproductive and endocrine systems may predispose women to a variety of diseases, the development of a particular disease dependent on the presence of additional genetic and environmental risk factors.

Interestingly, the direction of effect at some shared risk loci differed between EOC histotypes. The hepatocyte nuclear factor 1 beta *(HNF1B)* locus showed the same direction of effect between endometriosis and CCOC but was different between endometriosis and HGSOC consistent with published observations between CCOC and HGSOC (Natanzon *et al*, 2018). *HNF1B* is consistently highly expressed in CCOC but the promoter is methylated in HGSOC suggesting absence of *HNF1B* is critical for development of the HGSOC histotype (Natanzon *et al*., 2018; Ross-Adams *et al*, 2016), potentially against a common background of genetic liability to endometriosis. *HNF1B* is a transcription factor that plays a vital role in tissue development, and regulation of genes involved in cell cycle modulation, apoptosis, oxidative stress response and epithelial mesenchymal transition and dysregulation of these pathways may suggest a role for the microenvironment in tumor development (Suzuki *et al*, 2015; Yamaguchi *et al*, 2014). Similarly, SNPs in the *SYNE1* locus on chromosome 6 have the same direction of effect between endometriosis and CCOC but opposite direction of effect between endometriosis and LGSOC.

We provide evidence of functional mechanisms by which genetic variants associated with these diseases may be impacting noncoding regulatory elements that control the expression of genes that, when perturbed, increase risk of endometriosis and or ovarian cancer. Overall, many target genes shared between endometriosis and EOC differed between histotypes supporting evidence from other analyses in this study that different genes and gene pathways may contribute to the causal relationship between endometriosis and the different histotypes. *LINC00339*, located in the chromosome 1 risk region was associated with risk of endometriosis and HGSOC. The expression of *LINC00339* and nearby *CDC42* has been associated with endometriosis previously and *LINC00339* has been reported as the likely target gene (Fung *et al*, 2018; Powell *et al*, 2016). Masuda *et al* (2019) also reports that this same locus on chromosome 1 is associated with risk of both endometriosis and ovarian cancer. Methylation at a CpG site near *GREB1* in endometrium and blood is associated with increased risk for endometriosis, CCOC, ENOC, HGSOC and MOC. This association has been identified for endometriosis previously with functional studies yet to determine the molecular mechanisms contributing to disease risk (Fung *et al*, 2015; Mortlock *et al*, 2019). Transcription of *GREB1* splice variants has been associated with variants in this region in ovarian tissue (2020). This gene is expressed in EOC tumours with studies suggesting a reliance on *ESR1*/*GREB1* signalling (Hodgkinson *et al*, 2018; Laviolette *et al*, 2014).

Overlap with chromatin biofeatures in ovarian and endometriosis tissues also highlighted potential target genes. Risk SNPs in the *VEZT* region overlapped the putative bidirectional promoter for *VEZ*T and *FGD6*. The lead SNP from the bi-variate meta-analysis in the *VEZT* locus (rs7309252) was in LD (r^2^=0.99) with a SNP (rs6538618) overlapping 10 regulatory biofeatures. Expression of both *VEZT* and *FGD6* have been associated with endometriosis risk previously(Fung *et al*., 2018; Mortlock *et al*., 2020b). Similarly, the lead SNPs in the *FSHB* and *MLLT10* loci were in LD (r^2^>0.8) with SNPs overlapping 11 biofeatures in the promoter region of ADP Ribosylation Factor Like GTPase 14 Effector Protein (*ARL14EP*) and *MLLT10* respectively and were associated with methylation at nearby CpG sites. Risk SNPs associated with endometriosis, HGSOC and LMPSOC span follicle stimulating hormone (FSH) subunit B *(FSHB)* and nearby *ARL14EP. ARL14EP* is expressed by many tissue types and plays a role in the movement of major histocompatibility class II molecules along the actin cytoskeleton. *FSHB* is expressed in the pituitary gland and plays an important role regulating reproductive function. Variants in the 11p14.1 locus near *FSHB* have been significantly associated with multiple reproductive traits and diseases including PCOS, uterine fibroids, circulating sex hormone levels and menstrual cycle characteristics (Day *et al*, 2018; Day *et al*, 2015; Day *et al*, 2017; Gallagher *et al*, 2019; Laisk *et al*, 2018; Mbarek *et al*, 2016; Ruth *et al*, 2016; Sapkota *et al*., 2017). The lead SNP from the bi-variate meta-analysis, rs11031005, is in LD with a FSHB promoter polymorphism (rs10835638), and enhancer polymorphism (rs11031006) involved in regulating the *FSHB* gene transcription (Bohaczuk *et al*, 2021; Grigorova *et al*, 2008; Trevisan *et al*, 2019). The locus containing the histone lysine methyltransferase DOT1L Cofactor (*MLLT10*) was associated with endometriosis risk in a recent endometriosis GWAS (Rahmioglu *et al*., 2018). Studies have also linked ovarian cancer susceptibility and endometriosis risk to subtle variations in regulation at the *MLLT10* promoter region. SNPs in LD (r^2^>0.8) with the lead variant from the bivariate meta-analysis have been annotated to the promoter of *MLLT10* and have been associated with changes of expression of nearby genes *C10orf140, C10orf114* and *NEBL* in primary EOC tissues and changes in expression of *NEBL* in endometrium, suggesting this promoter may also have *cis* regulatory activity across the locus (Mortlock *et al*., 2020b; Pharoah *et al*, 2013).

This study uses a comprehensive range of statistical genetic approaches to build on existing evidence of an association between endometriosis and ovarian cancer using genetic data from the largest GWAS meta-analyses of endometriosis and EOC currently available. The power of this study to identify shared risk loci and target genes is however limited by the sample size of some of the less common EOC histotype cohorts such as LMPSOC. The identification of genetic relationships may also be limited by phenotypic annotation and heterogeneity between endometriosis cases affecting the endometriosis GWAS. Several studies have reported an association between endometriosis sub-phenotypes and risk of ovarian cancer, in particular endometriomas (Buis *et al*, 2013; Kobayashi *et al*, 2007; Kok *et al*., 2015; Saavalainen *et al*, 2018). More comprehensive phenotyping and molecular characterisation of endometriosis lesions could be used to test genetic associations between potential endometriosis subtypes and risk of certain EOC histotypes, for example testing if the association between endometriosis and CCOC is driven by endometriomas specifically.

In conclusion we find evidence of a strong genetic correlation and causal relationship between endometriosis and two ovarian cancer histotypes CCOC and ENOC and to a lesser extent HGSOC. Further investigation into shared genomic regions reveals different genetic variants, genes and pathways are likely contributing to the causal relationship with the different histotypes. Results add to our understanding of disease pathogenesis and yield genomic targets that may facilitate preventive pharmacological intervention by disrupting the link between endometriosis and ovarian cancer and promote targeted ovarian cancer screening in women with endometriosis.

## Methods

### Datasets

#### Epithelial Ovarian Cancer

Genetic data in the form of GWAS summary statistics were available from a 2017 GWAS meta-analysis for EOC and EOC histotypes conducted by Phelan *et al*. (2017). Summary statistics included the SNP RSID, effect allele, other allele, effect allele frequency, beta coefficient (be), standard error (SE) and p-value. The meta-analysis included 25,509 EOC cases and 40,941 controls of European ancestry with association statistics reported for 10,197,379 SNPs for overall EOC, high-grade serous ovarian cancer (HGSOC), low-grade serous ovarian cancer (LGSOC), low malignant potential serous ovarian cancer (LMPSOC), mucinous ovarian cancer (MOC), endometrioid ovarian cancer (ENOC) and clear cell ovarian cancer (CCOC). The number of cases and controls included in each histotype analysis are shown in Supplementary Table 11. Summary statistics were filtered to remove SNPs with an imputation quality *r*^2^ (OncoArray) score < 0.3 and SNPs with a minor allele frequency (MAF) < 0.01 leaving 10,197,379 SNPs for subsequent analysis.

#### Endometriosis

Endometriosis GWAS summary statistics were available from the 2017 GWAS meta-analysis for endometriosis conducted by Sapkota *et al*. (2017). Summary statistics include the SNP RSID, effect allele, other allele, beta coefficient (be), standard error (SE), effect allele frequency, and p-value. Only statistics generated from European cohorts was used in subsequent analyses including 14,949 cases and 190,715 controls. Summary statistics for 7,899,415 SNPs remained following removal of imputed genotypes with low imputation quality (<0.3 for minimac and <0.4 for IMPUTE2) and SNPs with a MAF < 0.01.

### Genetic Correlation and Mendelian Randomization

Linkage disequilibrium score regression (LDSC) was used to estimate the genetic correlation between endometriosis and ovarian cancer histotypes using the GWAS summary statistics (Bulik-Sullivan *et al*., 2015). LD scores were computed using 1000 Genomes European ancestry data as the independent variable in the LD Score regression and for the regression weights. In the absence of sample overlap between the datasets we also constrained the LD score regression intercept to reduce the standard error substantially. Mendelian randomization (MR) analyses were performed using the R package MendelianRandomization version 0.5.0 (Yavorska & Burgess, 2017). Inverse-variance weighted (IVW)(Burgess *et al*., 2013) regression was used for primary analysis and weighted median (Bowden *et al*., 2016) and MR-Egger regression(Bowden *et al*., 2015) for sensitivity analyses. Associations were declared significant if both IVW and weighted median analyses yield P-values < 0.05 and the direction of the effect size estimates (odds ratios) were consistent across IVW, weighted median, MR-Egger regression approaches. Of the 27 independent, genome-wide significant endometriosis lead risk SNPs (Rahmioglu *et al*., 2018), two multiallelic SNPs (rs484686 and rs4762173) were removed, leaving 25 SNPs in the instrument for genetic liability to endometriosis.

### SNP-based Association Analysis

#### Cross-trait Meta-analysis

To identify risk loci associated with both traits we conducted a bivariate meta-analysis using two different approaches, MetABF (Trochet *et al*., 2019) and Han and Eskin random-effects model (RE2C) (Han & Eskin, 2011; Lee *et al*., 2017). MetABF is a method to meta-analyse genome-wide association studies using approximate Bayes Factors. Beta coefficients (effect sizes) and SEs from the endometriosis dataset were used as input for MetABF alongside beta coefficients and SEs from ovarian cancer histotypes. Each of the six ovarian cancer histotypes were meta-analysed with endometriosis separately. Correlation in effect sizes was modeled using both an independent and fixed effect model and the prior parameter for the variance in effect sizes, sigma, was set to 0.1 allowing effect sizes to be small, characteristic of complex diseases. Significantly associated SNPs were defined by log_10_ ABF > 4 in either the fixed or independent model and at least nominal significance in the single trait GWAS (P-value<0.05).

To validate results, we also performed a routine fixed effects meta-analysis and a modified random effects meta-analysis on the same data using RE2C. RE2C is designed to integrate the effects while accounting for the heterogeneity between studies. A SNP was deemed significantly associated with both traits in the bivariate meta-analysis if it met a fixed (Cochran’s Q statistic P-value >0.05) or random (Cochran’s Q statistics’s P-value <0.05) effect threshold of P-value < 5×10^−8^, and was at least nominally significant in the single trait GWAS meta-analysis (P-value<0.05) and had no significant heterogeneity. SNPs found to be significantly associated with both traits and passing thresholds for both Metabf and RE2C were fine mapped using FUMA (Watanabe *et al*., 2017) to identify independent signals to identify independent signals at *r*^2^ < 0.6.

#### Colocalization Analyses

GWAS-pw (Pickrell *et al*., 2016) was used to estimate the probability that in a given genomic region the same variant underlies the association with both traits. The genome is split into 1703 non-overlapping regions using linkage disequilibrium blocks (Berisa & Pickrell, 2015) and following Giambartolomei *et al* (2014) the software estimates the probability, using an empirical Bayes approach, that a given genomic region either 1) contains a genetic variant that influences the first trait, 2) contains a genetic variant that influences the second trait, 3) contains a genetic variant that influences both traits (posterior probability of association, PPA3), or 4) contains both a genetic variant that influences the first trait and a separate genetic variant that influences the second trait (PPA4). This was performed using the endometriosis dataset and each ovarian cancer histotype. Regions with a PPA3/4>0.5 were considered to have evidence of a shared causal variant and independent causal variants respectively.

### Gene-based Association Analysis

The fastBat function in GCTA (Bakshi *et al*., 2016) was used to perform a gene-based association analysis for endometriosis and each ovarian cancer histotype using GWAS summary statistics from each trait. A total of 20,439 genes (hg19) were tested for each disease using an LD cutoff of 0.9 and no SNPs outside defined gene boundaries. Genes in the top 1% and 5% associated with each EOC histotype that overlap with the top 1% and 5% of genes associated with endometriosis were tested for enrichment in pathways using over-representation analysis in WebGestalt (Liao *et al*., 2019).

### Functional Annotation

The GENE2FUNC option in FUMA (Watanabe *et al*., 2017) was also used to investigate the expression of nearby (within 10kb) genes across tissues and enrichment of functional and biological pathways.

#### H3K27ac ChIP-seq and ATAC-seq

H3K27ac ChIP-seq data have been previously described (Coetzee *et al*, 2015a; Corona *et al*, 2020b; Jones *et al*, 2020) with the exception of the endometrioma and endometrioma stroma specimens, which were profiled in parallel with the tumors reported in Corona *et al*. (2020b).

ATAC-seq data were generated for primary tissues collected, with informed consent, as part of the Gynecologic Tissue Bank at Cedars-Sinai Medical Center. Tumors were OCT embedded and H&Es reviewed to confirm the diagnosis. Epithelial-rich regions of flash frozen tumors were biopsied using a 5mm biopsy punch. Primary fallopian tube tissues were subjected to enzymatic digest, as previously described (Fotheringham *et al*, 2011; Karst & Drapkin, 2012) and the epithelial-enriched cells frozen down for ATAC-seq. In one instance, fallopian epithelial cells were frozen prior to profiling. ATAC-seq was performed by Active Motif.

##### Peak calling

Peak calling of H3K27ac ChIP-seq profiles was performed using the ENCODE pipeline (v1.2.2) with p-val_thresh = 1e-09, reference genome hg38 and other default parameters. If two technical replicates were used, we selected the overlap peak set of the true replicates (rep1_rep2.overlap.bfilt.narrowPeak.gz) as the ‘sample peak set’, otherwise, we used the overlap peak set of the pseudo replicates (rep1-pr.overlap.bfilt.narrowPeak.gz). Quality control metrics (number of unique mapped reads, cross-correlation metrics, IDR) were checked for each technical replicate (Supplementary Table 12).

##### Donor peak set

Two or more sample peak sets are combined if they belong to the same donor, resulting in a set of non-overlapping peaks. First, individual peak scores (-log10(p-value)) are normalized to a ‘score per million’ (SPM) dividing the peak score, by the sum of all peak scores within the sample, divided by a factor of a million. Then, all peaks from the same donor are merged and sorted by SPM. Iteratively, a peak is removed if it overlaps another peak with a higher SPM, resulting in a set of non-overlapping peaks that represent a donor.

##### Consensus peak set

A set of sample/donor peak sets will be combined when they are part of the same experiment, tissue type and sample type triage. Similarly to the donor peak set procedure, peaks are merged, sorted and iteratively removed, based on the SPM. Then, the remaining non-overlapping peaks are tagged as reproducible if they overlap peaks with an SPM > 5 in two or more samples. Non-reproducible peaks, peaks that are within repeat mask regions and peaks in the “Y” chromosome are removed, leaving a set of reproducible non-overlapping peaks as the ‘consensus peak set’. Consensus peak sets are then transformed from hg38 to hg19 using *liftOver* with parameteres -bedPlus=6, the hg38toHg19.over.chain.gz from UCSC chain files and other default parameters. On average, each consensus peak set has 125.2 (0.28%) genomic locations that could not be mapped (range = [8,350]).

#### Summary-data-based Mendelian randomization (SMR)

Summary-data-based Mendelian randomization (SMR) is used to assess the pleiotropic and causal association between genetic variants, gene expression/methylation level and risk of disease (Zhu *et al*., 2016). SMR has been conducted using summary statistics from Sapkota *et al*. (2017) and endometrial and blood eQTLs and mQTLs (Fung *et al*., 2017; McRae *et al*., 2018; Mortlock *et al*., 2020a; Mortlock *et al*., 2019). Using these published results, we searched for any significant SMR associations in loci associated (log_10_ ABF > 4 in the cross-trait MetABF analysis, P-value < 5×10^−8^ in the cross-trait RE2C analysis and P-value < 0.05 in each single trait meta-analysis) with both endometriosis and ovarian cancer from the bivariate meta-analysis. We also conducted SMR on each ovarian cancer histotype by integrating the GWAS meta-analysis summary data from Phelan *et al*. (2017) and summary eQTL data from endometrium (Fung *et al*., 2017; Mortlock *et al*., 2020a), eQTLGen (Võsa *et al*., 2018) and GTEx ovary and uterus (Consortium *et al*., 2017). Associations were considered significant if they had a P_SMR_<0.05/(number of genes tested) and a *P*_HEIDI_ > 0.05/(number of genes passing the SMR test). Of note, associations with P_SMR_<0.05 and a *P*_HEIDI_ > 0.05 were also considered due to power limitations in existing datasets. In the absence of multiple testing correction these would require future validation in larger datasets or functional studies. The heterogeneity in dependent instruments (HEIDI) test considers the pattern of risk associations using all the SNPs that are significantly associated with gene expression in a region and evaluates the null hypothesis that there is a single variant affecting gene expression/methylation and disease risk and the alternative hypothesis that there are distinct variants associated with expression/methylation and disease.

## Data Availability

This study includes no data deposited in external repositories.

## Data Availability

This study includes no data deposited in external repositories.

## Acknowledgements

We would like to thank the research participants and employees of 23andMe for making this work possible. We would also like to acknowledge the International Endometriosis Genetics Consortium and Ovarian Cancer Association Consortium for their contributions generating the GWAS datasets and data access. We are also grateful to the thousands of patients who donated the specimens that enable this research to happen. For acknowledgements for the endometriosis meta-analysis please see Sapkota *et al*. (2017). For acknowledgements for the ovarian cancer meta-analysis please see Phelan *et al*. (2017).

## Funding

This work was supported by the National Health and Medical Research Council of Australia (GNT1026033, GNT1105321, GNT1147846, Investigator Grant 1177194 to G.W.M and Medical Research Future Fund Research Grant MRF1199785 to S.M) and National Institutes of Health (R01CA193910, R01CA204954, R01CA211707, R01CA251555). S.P.K is supported by a United Kingdom Research and Innovation Future Leaders Fellowship (MR/T043202/1). K.L. is supported by a Liz Tilberis Early Career Award (599175) and a Program Project Development (373356) from the Ovarian Cancer Research Alliance, plus a Research Scholar’s Grant from the American Society (134005). For funding details of the endometriosis meta-analysis please see Sapkota *et al*. (2017). For funding details of the ovarian cancer meta-analysis please see Phelan *et al*. (2017).

## Author contributions

S.M, S.P.K, K.L, G.W.M and P.P designed the study with input from the other authors. Data analysed in this study was generated by the Ovarian Cancer Association Consortium and International Endometriosis Genetics Consortium. S.M, and S.P.K ran additional quality control and filtering of GWAS datasets. J-H.S, M.L.F, S.A.G, M.T.S, R.L, C.W, I.C, B.Y.K, P.A.W.R and B.J.R contributed to specimen collection and data generation. Data analysis was performed by S.M, S.P.K, R.I.C and P.F.K which was interpreted by all authors. S.M, S.P.K and K.L drafted the report with input from all other authors. The final manuscript has been critically revised and approved by all authors.

## Conflict of interest

The authors declare that they have no conflict of interest.

## Notes

### Competing Interest Statement

The authors have declared no competing interest.

