## Supplementary Figure for "A multi-level investigation of the genetic relationship between endometriosis and ovarian cancer histotypes"

### Supplementary Figures

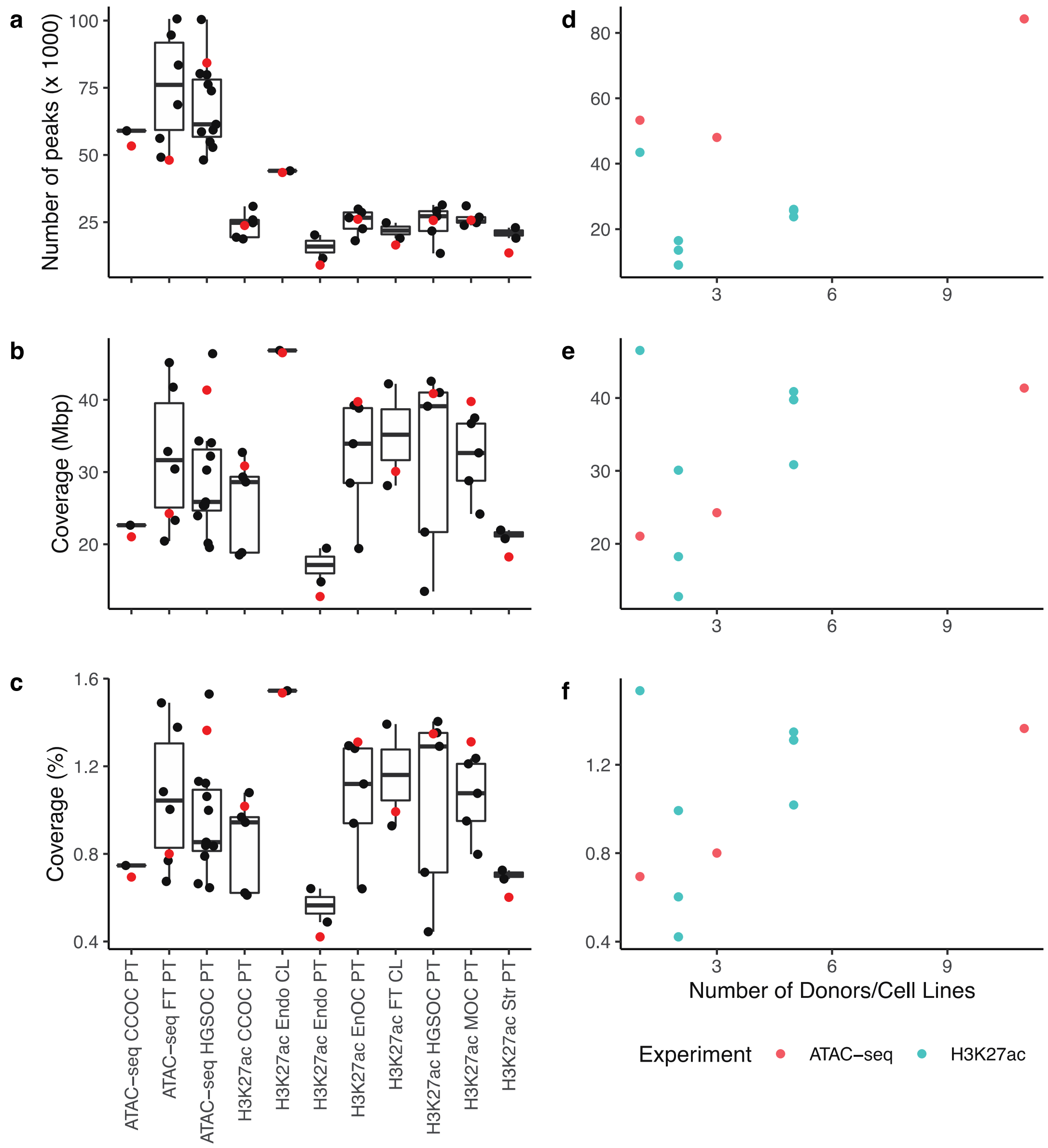


**Figure S1.** (a) Number of peaks and (b) genome coverage in Mbp and (c) genome coverage in percentage for of sample peak sets (black dots) and consensus peak sets (red dots). (d) Number of peaks (e) and genome coverage in Mbp and (g) genome coverage in percentage for consensus peak sets as a function of the number of donors.

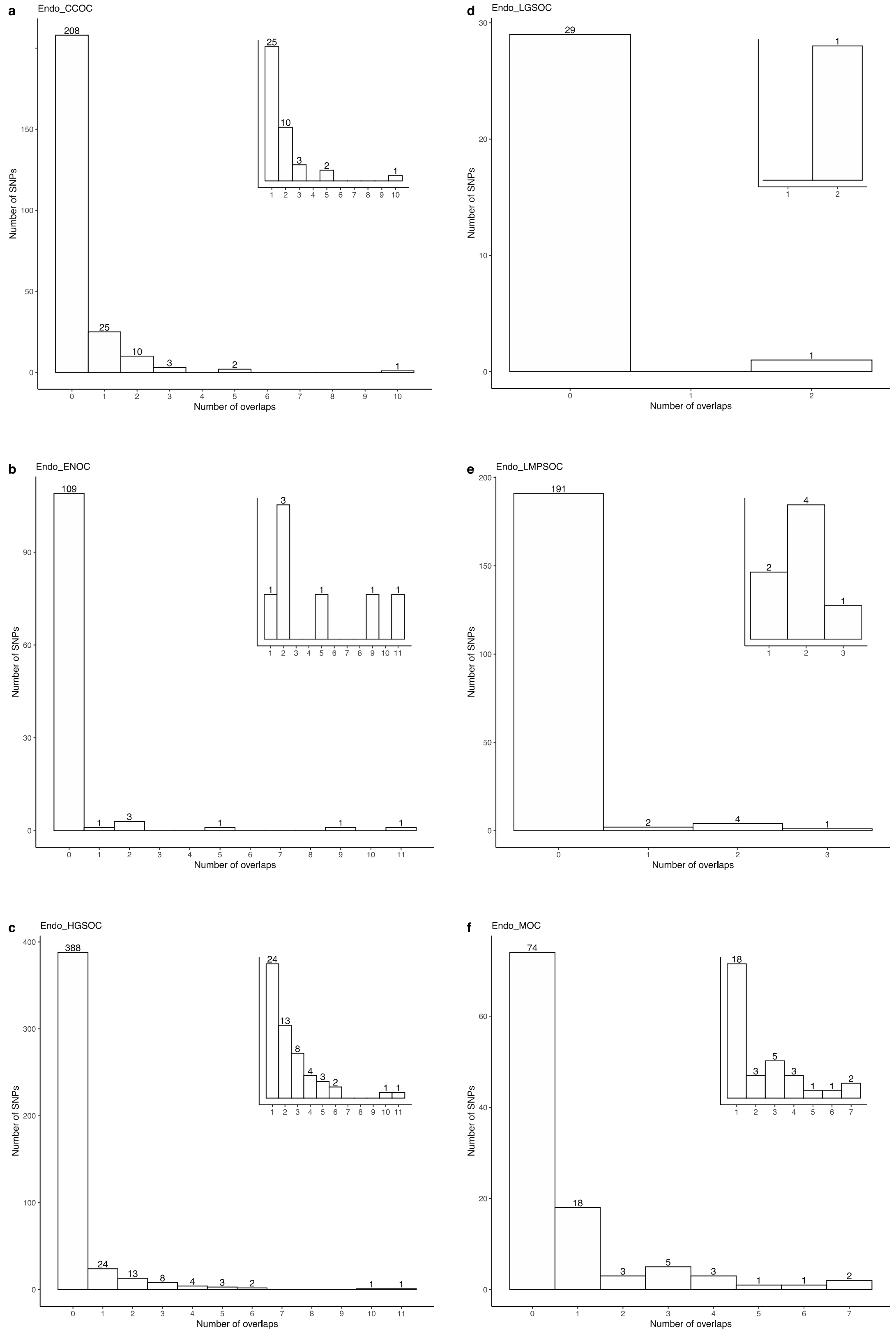


**Figure S2.** Histogram of number of SNPs that overlap *n* biofeatures for (a) CCOC, (b) EnOC, (c) HGSOC, (d) LGSOC, (e) LMPSOC and (f) MOC. Inset histograms show the number of overlaps ≥ 1.

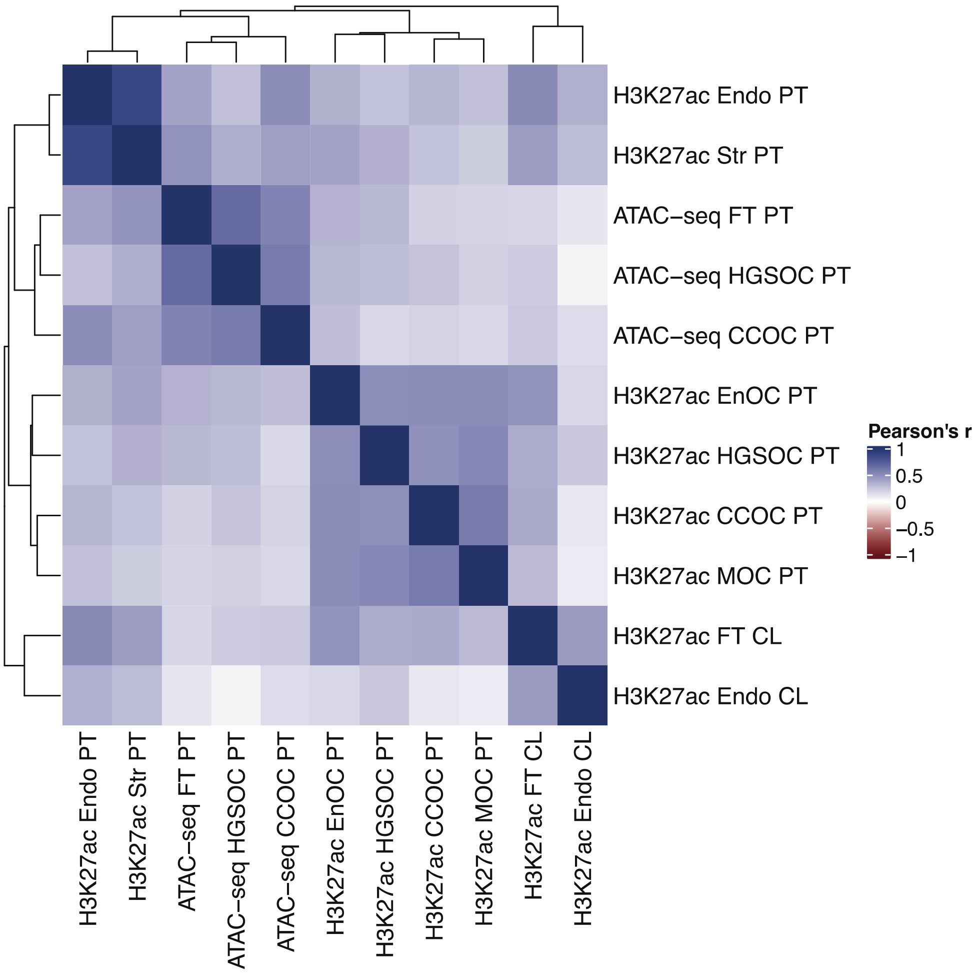


**Figure S3.** Pearson's correlation coefficient between biofeatures based on the overlap with endometriosis/OC risk SNPs.

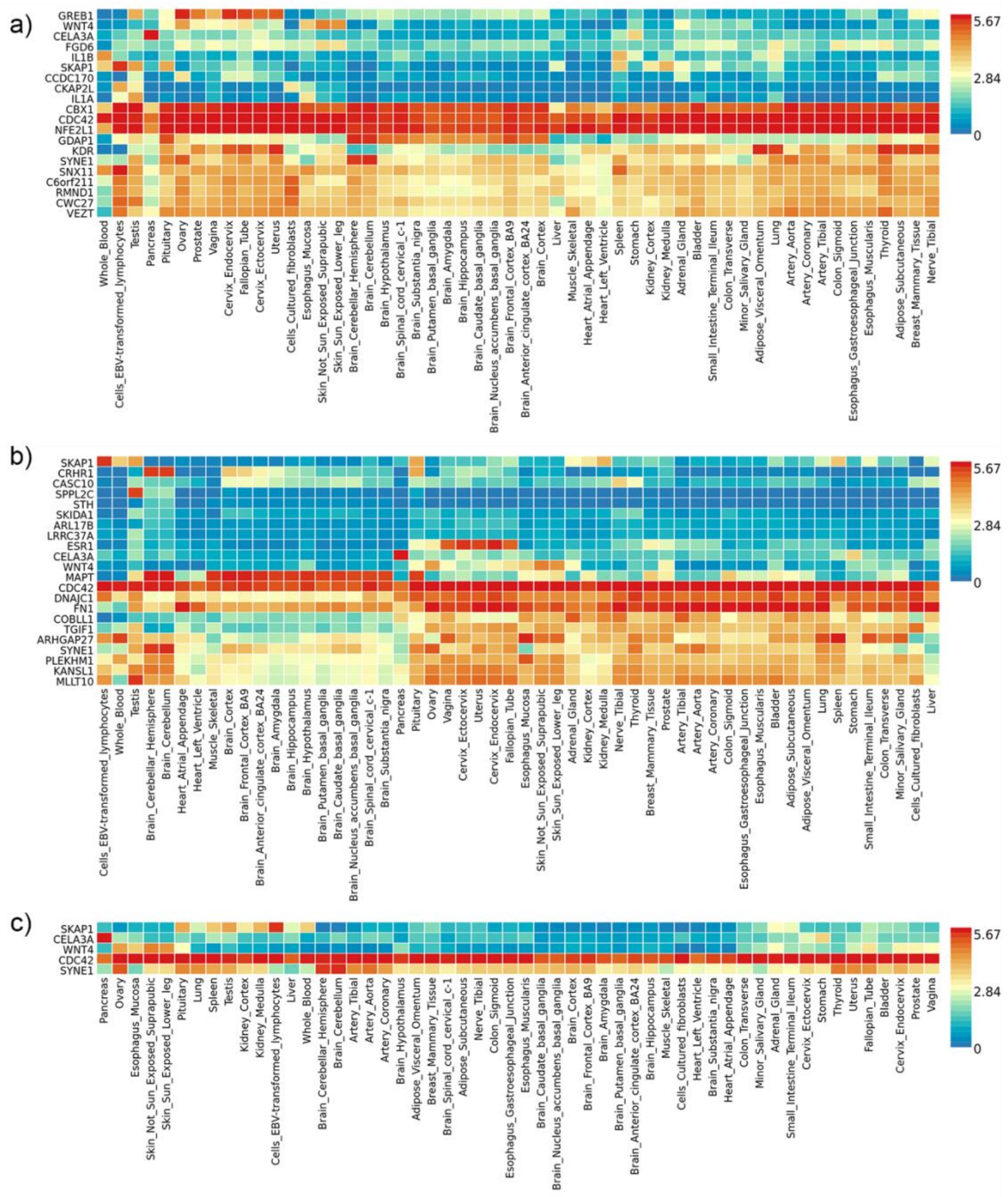


**Figure S4.** Heatmap, generated in FUMA, of genes annotated to SNPs significantly associated with endometriosis plus (a) CCOC, (b) HGSOC and (c) all three, expressed across 58 tissues from GTEx.

### International Endometriosis Genetics Consortium

Yadav Sapkota[1,2], Valgerdur Steinthorsdottir[3], Andrew P. Morris[4,5], Amelie Fassbender[6,7], Nilufer Rahmioglu[5], Immaculata De Vivo[8,9], Julie E. Buring[8,10], Futao Zhang[11], Todd L. Edwards[12], Sarah Jones[13], Dorien O[6,7], Daniëlle Peterse[6,7], Kathryn M. Rexrode[8,10], Paul M. Ridker[8,10], Andrew J. Schork[14,15], Stuart MacGregor[1], Nicholas G. Martin[1], Christian M. Becker[16], Sosuke Adachi[17], Kosuke Yoshihara[17], Takayuki Enomoto[17], Atsushi Takahashi[18], Yoichiro Kamatani[18], Koichi Matsuda[19], Michiaki Kubo[18], Gudmar Thorleifsson[3], Reynir T. Geirsson[20,21], Unnur Thorsteinsdottir[3,21], Leanne M. Wallace[1,11], iPSYCH-SSI-Broad Groupw, Jian Yang[11], Digna R. Velez Edwards[22], Mette Nyegaard[23,24], Siew-Kee Low[18], Krina T. Zondervan[5,16], Stacey A. Missmer[8,9], Thomas D'Hooghe[6,7,25], Grant W. Montgomery[1,11], Daniel I. Chasman[8,10], Kari Stefansson[3,21], Joyce Y. Tung[26] & Dale R. Nyholt[1,27].

[1]Department of Genetics and Computational Biology, QIMR Berghofer Medical Research Institute, Brisbane, Queensland 4006, Australia. [2]Department of Epidemiology and Cancer Control, St. Jude Children's Research Hospital, Memphis, Tennessee 38105, USA. [3] deCODE Genetics/Amgen, 101 Reykjavik, Iceland. [4]Department of Biostatistics, University of Liverpool, Liverpool L69 3GL, UK. [5]Wellcome Trust Centre for Human Genetics, University of Oxford, Oxford OX3 7BN, UK. [6] KULeuven, Department of Development and Regeneration, Organ systems, 3000 Leuven, Belgium. [7]Department of Obstetrics and Gynaecology, Leuven University Fertility Centre, University Hospital Leuven, 3000 Leuven, Belgium. [8]Harvard T.H. Chan School of Public Health, Boston, Massachusetts 02115, USA. [9] Channing Division of Network Medicine, Department of Medicine, Brigham and Women's Hospital and Harvard Medical School, Boston, Massachusetts 02115, USA. [10]Division of Preventive Medicine, Brigham and Women's Hospital, Boston, Massachusetts 02215, USA. [11] Institute for Molecular Bioscience, The University of Queensland, Brisbane, Queensland 4072, Australia. [12] Institute of Medicine and Public Health, Vanderbilt University Medical Center, Nashville, Tennessee 37203, USA. [13] Vanderbilt Genetics Institute, Division of Epidemiology, Institute of Medicine and Public Health, Department of Medicine, Vanderbilt University Medical Center, Nashville, Tennessee 37203, USA. [14] Cognitive Science Department, University of California, San Diego, La Jolla, California 92093, USA. [15] Institute of Biological Psychiatry, Mental Health Centre Sct. Hans, Copenhagen University Hospital, DK-2100 Copenhagen, Denmark. [16] Endometriosis CaRe Centre, Nuffield Dept of Obstetrics & Gynaecology, University of Oxford, John Radcliffe Hospital, Oxford OX3 9DU, UK. [17]Department of Obstetrics and Gynecology, Niigata University Graduate School of Medical and Dental Sciences, Niigata 950-2181, Japan. [18] Center for Integrative Medical Sciences, RIKEN, Yokohama 230-0045, Japan. [19] Institute of Medical Sciences, The University of Tokyo, Tokyo 108-8639, Japan. [20]Department of Obstetrics and Gynecology, Landspitali University Hospital, 101 Reykjavik, Iceland. [21] Faculty of Medicine, School of Health Sciences, University of Iceland, 101 Reykjavik, Iceland. [22]Vanderbilt Genetics Institute, Vanderbilt Epidemiology Center, Institute of Medicine and Public Health, Department of Obstetrics and Gynecology, Vanderbilt University Medical Center, Nashville, Tennessee 37203, USA. [23]Department of Biomedicine, Aarhus University, DK8000 Aarhus, Denmark. [24] iPSYCH, The Lundbeck Foundation Initiative for Integrative Psychiatric Research, DK-2100 Copenhagen, Denmark. [25]Global Medical Affairs Fertility, Research and Development, Merck KGaA, Darmstadt, Germany. [26] 23andMe, Inc., 899 W. Evelyn Avenue, Mountain View, California 94041, USA. [27] Institute of Health and Biomedical Innovation, Queensland University of Technology, Queensland 4059, Australia.

### Ovarian Cancer Association Consortium

Hoda Anton-Culver[1], Elisa V. Bandera[2], Susana N Banerjee[3], Javier Benitez[4], Andrew Berchuck[5], Line Bjorge[6], Ingrid A. Boere[7], James D. Brenton[8], Ralf Butzow[9], Ian Campbell[10], Kexin Chen[11], Georgia Chenevix-Trench[12], Linda S. Cook[13], Daniel W. Cramer[14], Anna deFazio[15], Jennifer A. Doherty[16], Thilo Dörk[17], Diana M. Eccles[18], Peter A. Fasching[19], Renée T. Fortner[20], Rosalind Glasspool[21], Ellen L. Goode[22], Marc T. Goodman[23], Jacek Gronwald[24], Claus K. Høgdall[25], Estrid Høgdall[26], Chad Hamilton[27], Holly R. Harris[28], Florian Heitz[29], Michelle A.T. Hildebrandt[30], Akira Hirasawa[31], Antoinette Hollestelle[7], David G. Huntsman[32], Issei Imoto[33], Beth Y. Karlan[34], Linda E. Kelemen[35], Lambertus A. Kiemeney[36], Susanne K. Kjaer[26], Anita Koushik[37], Mieke Kriege[7], Björg Kristjansdottir[38], Jolanta Kupryjanczyk[39], Diether Lambrechts[40], Nhu D. Le[41], Douglas A. Levine[42], Keitaro Matsuo[43], G Larry Maxwell[27], Taymaa May[44], Iain A. McNeish[45], Usha Menon[46], Roger L. Milne[47], Francesmary Modugno[48], Alvaro N. Monteiro[49], Patricia G. Moorman[50], Kirsten B. Moysich[51], Heli Nevanlinna[52], Sara H. Olson[53], Håkan Olsson[54], Sue K. Park[55], Celeste L. Pearce[56], Tanja Pejovic[57], Malcolm C. Pike[53], Susan J. Ramus[58], Elio Riboli[59], Marjorie J. Riggan[5], Harvey A. Risch[60], Cristina Rodriguez-Antona[4], Isabelle Romieu[61], Dale P. Sandler[62], Joellen M. Schildkraut[63], V. Wendy Setiawan[64], Kang Shan[65], Nadeem Siddiqui[66], Weiva Sieh[67], Meir Stampfer[68], Karin Sundfeldt[38], Rebecca Sutphen[69], Anthony J. Swerdlow[70], Soo Hwang Teo[71], Kathryn L. Terry[14], Shelley S. Tworoger[49], Digna Velez Edwards[72], Roel C.H. Vermeulen[73], Penelope M. Webb[74], Nicolas Wentzensen[75], Emily White[76], Walter Willett[77], Alicja Wolk[78], Yin-Ling Woo[79], Anna H. Wu[64], Li Yan[80], Drakoulis Yannoukakos[81], Wei Zheng[82]

[1] Department of Medicine, Genetic Epidemiology Research Institute, University of California Irvine, Irvine, CA, USA. [2] Cancer Prevention and Control Program, Rutgers Cancer Institute of New Jersey, New Brunswick, NJ, USA. [3] Gynaecology Unit, Royal Marsden Hospital, London, UK. [4] Biomedical Network on Rare Diseases (CIBERER), Madrid, Spain. [5] Department of Gynecologic Oncology, Duke University Hospital, Durham, NC, USA. [6] Department of Obstetrics and Gynecology, Haukeland University Hospital, Bergen, Norway. [7] Department of Medical Oncology, Erasmus MC Cancer Institute, Rotterdam, The Netherlands. [8] Cancer Research UK Cambridge Institute, University of Cambridge, Cambridge, UK. [9] Department of Pathology, Helsinki University Hospital, University of Helsinki, Helsinki, Finland. [10] Peter MacCallum Cancer Center, Melbourne, Victoria, Australia. [11] Department of Epidemiology, Tianjin Medical University Cancer Institute and Hospital, Tianjin, China. [12] Department of Genetics and Computational Biology, QIMR Berghofer Medical Research Institute, Brisbane, Queensland, Australia. [13] University of New Mexico Health Sciences Center, University of New Mexico, Albuquerque, NM, USA. [14] Department of Epidemiology, Harvard T.H. Chan School of Public Health, Boston, MA, USA. [15] Centre for Cancer Research, The Westmead Institute for Medical Research, The University of Sydney, Sydney, New South Wales, Australia. [16] Huntsman Cancer Institute, Department of Population Health Sciences, University of Utah, Salt Lake City, UT, USA. [17] Gynaecology Research Unit, Hannover Medical School, Hannover, Germany. [18] Faculty of Medicine, University of Southampton, Southampton, UK. [19] David Geffen School of Medicine, Department of Medicine Division of Hematology and Oncology, University of California at Los Angeles, Los Angeles, CA, USA. [20] Division of Cancer Epidemiology, German Cancer Research Center (DKFZ), Heidelberg, Germany. [21] Department of Medical Oncology, Beatson West of Scotland Cancer Centre and University of Glasgow, Glasgow, UK. [22] Department of Health Science Research, Division of Epidemiology, Mayo Clinic, Rochester, MN, USA. [23] Samuel Oschin Comprehensive Cancer Institute, Cancer Prevention

and Genetics Program, Cedars-Sinai Medical Center, Los Angeles, CA, USA. [24] Department of Genetics and Pathology, Pomeranian Medical University, Szczecin, Poland. [25] Department of Gynaecology, Rigshospitalet, University of Copenhagen, Copenhagen, Denmark. [26] Department of Virus, Lifestyle and Genes, Danish Cancer Society Research Center, Copenhagen, Denmark. [27] Gynecologic Cancer Center of Excellence, Inova Schar Cancer Institute, Falls Church, VA, USA. [28] Program in Epidemiology, Division of Public Health Sciences, Fred Hutchinson Cancer Research Center, Seattle, WA, USA. [29] Humboldt-UniversitŠ t zu Berlin, and Berlin Institute of Health, Department for Gynecology with the Center for Oncologic Surgery CharitŽ Campus Virchow-Klinikum, CharitŽ Ð UniversitŠ tsmedizin Berlin, corporate member of Freie UniversitŠ t Berlin, Berlin, Germany. [30] Department of Epidemiology, University of Texas MD Anderson Cancer Center, Houston, TX, USA. [31] Department of Clinical Genomic Medicine, Graduate School of Medicine, Dentistry and Pharmaceutical Sciences, Okayama University, Okayama, Japan. [32] British Columbia's Ovarian Cancer Research (OVCARE) Program, BC Cancer, Vancouver General Hospital, and University of British Columbia, Vancouver, BC, Canada. [33] Department of Human Genetics, Graduate School of Biomedical Sciences, Tokushima University, Tokushima, Japan. [34] David Geffen School of Medicine, Department of Obstetrics and Gynecology, University of California at Los Angeles, Los Angeles, CA, USA. [35] Hollings Cancer Center, Medical University of South Carolina, Charleston, SC, USA. [36] Radboud Institute for Health Sciences, Radboud University Medical Center, Nijmegen, The Netherlands. [37] CHUM Research Centre (CRCHUM), MontrŽ al, QC, Canada. [38] Department of Obstetrics and Gynecology, Sahglrenska Cancer Center, Inst Clinical Scienses, Sahlgrenska Academy at University of Gothenburg, Gothenburg, Sweden. [39] Department of Pathology and Laboratory Diagnostics, Maria Sklodowska-Curie National Research Institute of Oncology, Warsaw, Poland. [40] VIB Center for Cancer Biology, Leuven, Belgium. [41] Cancer Control Research, BC Cancer, Vancouver, BC, Canada. [42] Gynecology Service, Department of Surgery, Memorial Sloan Kettering Cancer Center, New York, NY, USA. [43] Division of Cancer Epidemiology and Prevention, Aichi Cancer Center Research Institute, Nagoya, Japan. [44] Division of Gynecologic Oncology, University Health Network, Princess Margaret Hospital, Toronto, Ontario, Canada. [45] Division of Cancer and Ovarian Cancer Action Research Centre, Department Surgery & Cancer, Imperial College London, London, UK. [46] Institute of Clinical Trials & Methodology, University College London, London, UK. [47] Cancer Epidemiology Division, Cancer Council Victoria, Melbourne, Victoria, Australia. [48] Womens Cancer Research Center, Magee-Womens Research Institute and Hillman Cancer Center, Pittsburgh, PA, USA. [49] Department of Cancer Epidemiology, Moffitt Cancer Center, Tampa, FL, USA. [50] Department of Community and Family Medicine, Duke University Hospital, Durham, NC, USA. [51] Division of Cancer Prevention and Control, Roswell Park Cancer Institute, Buffalo, NY, USA. [52] Department of Obstetrics and Gynecology, Helsinki University Hospital, University of Helsinki, Helsinki, Finland. [53] Department of Epidemiology and Biostatistics, Memorial Sloan-Kettering Cancer Center, New York, NY, USA. [54] Department of Cancer Epidemiology, Clinical Sciences, Lund University, Lund, Sweden. [55] Department of Preventive Medicine, Seoul National University College of Medicine, Seoul, Korea. [56] Department of Epidemiology, University of Michigan School of Public Health, Ann Arbor, MI, USA. [57] Department of Obstetrics and Gynecology, Oregon Health & Science University, Portland, OR, USA. [58] School of Women's and Children's Health, Faculty of Medicine, University of NSW Sydney, Sydney, New South Wales, Australia. [59] Imperial College London, London, UK. [60] Chronic Disease Epidemiology, Yale School of Public Health, New Haven, CT, USA. [61] Nutrition and Metabolism Section, International Agency for Research on Cancer (IARC-WHO), Lyon, France. [62] Epidemiology Branch, National Institute of Environmental Health Sciences, NIH, Research Triangle Park,

NC, USA. [63] Department of Epidemiology, Rollins School of Public Health, Emory University, Atlanta, GA, USA. [64] Department of Preventive Medicine, Keck School of Medicine, University of Southern California, Los Angeles, CA, USA. [65] Department of Obstetrics and Gynaecology, Hebei Medical University, Fourth Hospital, Shijiazhuang, China. [66] Department of Gynaecological Oncology, Glasgow Royal Infirmary, Glasgow, UK. [67] Department of Population Health Science and Policy, Icahn School of Medicine at Mount Sinai, New York, NY, USA. [68] Channing Division of Network Medicine, Department of Medicine, Brigham and Women's Hospital and Harvard Medical School, Boston, MA, USA. [69] Epidemiology Center, College of Medicine, University of South Florida, Tampa, FL, USA. [70] Division of Genetics and Epidemiology, The Institute of Cancer Research, London, UK. [71] Breast Cancer Research Programme, Cancer Research Malaysia, Subang Jaya, Selangor, Malaysia. [72] Division of Quantitative Sciences, Department of Obstetrics and Gynecology, Department of Biomedical Sciences, Women's Health Research, Vanderbilt University Medical Center, Nashville, TN, USA. [73] Julius Center for Health Sciences and Primary Care, University Utrecht, UMC Utrecht, Utrecht, The Netherlands. [74] Population Health Department, QIMR Berghofer Medical Research Institute, Brisbane, Queensland, Australia. [75] Division of Cancer Epidemiology and Genetics, National Cancer Institute, Bethesda, MD, USA. [76] Fred Hutchinson Cancer Research Center, Seattle, WA, USA. [77] Department of Nutrition, Harvard T.H. Chan School of Public Health, Boston, MA, USA. [78] Institute of Environmental Medicine, Karolinska Institutet, Stockholm, Sweden. [79] Department of Obstetrics and Gynaecology, University of Malaya Medical Centre, University of Malaya, Kuala Lumpur, Malaysia. [80] Department of Molecular Biology, Hebei Medical University, Fourth Hospital, Shijiazhuang, China. [81] Molecular Diagnostics Laboratory, INRASTES, National Centre for Scientific Research "Demokritos", Athens, Greece. [82] Division of Epidemiology, Department of Medicine, Vanderbilt Epidemiology Center, Vanderbilt-Ingram Cancer Center, Vanderbilt University School of Medicine, Nashville, TN, USA
